# Dynamic regulatory elements in single-cell multimodal data capture autoimmune disease heritability and implicate key immune cell states

**DOI:** 10.1101/2023.02.24.23286364

**Authors:** Anika Gupta, Kathryn Weinand, Aparna Nathan, Saori Sakaue, Accelerating Medicines Partnership RA/SLE Program and Network, Laura Donlin, Kevin Wei, Alkes L. Price, Tiffany Amariuta, Soumya Raychaudhuri

**Affiliations:** Center for Data Sciences, Brigham and Women’s Hospital and Harvard Medical School, Boston, MA, USA; Division of Rheumatology, Inflammation, and Immunity, Department of Medicine, Brigham and Women’s Hospital and Harvard Medical School, Boston, MA, USA; Division of Genetics, Department of Medicine, Brigham and Women’s Hospital and Harvard Medical School, Boston, MA, USA; Program in Medical and Population Genetics, Broad Institute of MIT and Harvard, Cambridge, MA, USA; Department of Biomedical Informatics, Harvard Medical School, Boston, MA, USA; Hospital for Special Surgery, New York, NY, USA; Weill Cornell Medicine, New York, NY, USA; Department of Epidemiology, Harvard T.H. Chan School of Public Health, Boston, MA, USA; Department of Biostatistics, Harvard T.H. Chan School of Public Health, Boston, MA, USA; Halıcıoğlu Data Science Institute, University of California San Diego, La Jolla, CA, USA; Department of Medicine, University of California San Diego, La Jolla, CA, USA

**Author notes:** Correspondence should be addressed to: Soumya Raychaudhuri, 77 Avenue Louis Pasteur, Harvard, New Research Building, Suite 250D Boston, MA 02115, USA., Tiffany Amariuta, 3180 Voigt Dr, UCSD, Franklin Antonio Hall, Office 2104 La Jolla, CA 92093, USA.

## Abstract

In autoimmune diseases such as rheumatoid arthritis (RA), the immune system attacks host tissues^1-3^. Developing a precise understanding of the fine-grained cell states that mediate the genetics of autoimmunity is critical to uncover causal disease mechanisms and develop potentially curative therapies. We leveraged multimodal single-nucleus (sn) RNA-seq and ATAC-seq data across 28,674 cells from the inflamed synovium of 12 donors with arthritis to identify accessible regions of chromatin associated with gene expression patterns that reflect cell states. For 12 autoimmune diseases, we discovered that cell-state-dependent (“dynamic”) peaks in immune cell types disproportionately captured heritability, compared to cell-state-invariant (“cs-invariant”) peaks. These dynamic peaks marked regulatory elements associated with T peripheral helper, regulatory T, dendritic, and STAT1^+^CXCL10^+^ myeloid cell states. We argue that dynamic regulatory elements can help identify precise cell states enriched for disease-critical genetic variation.

---

Recent single-cell sequencing studies that probe inflamed tissues from patients with autoimmune diseases have identified expanded functional cell states. Expanded T cell states include CD4^+^ T peripheral helper (Tph) cells in RA^4-6^, ulcerative colitis (UC)^7^, and Celiac disease (celiac)^8^; clonally distinct T follicular helper (Tfh) cells in systemic lupus erythematosus (SLE)^9^; GZMK^+^CD8^+^ T cells in RA^10^; and CD39^+^CCR6^+^ and CD39^+^CD4^+^ Type 17 helper cells in Crohn’s disease (CD)^11^. Interferon (IFN)-imprinted naive and IFN-stimulated B cells are expanded in UC^7^ and lupus nephritis^12^, respectively. Single-cell studies have also identified myeloid-derived cell states associated with autoimmunity, including CD14^+^ dendritic cells (DCs) in psoriasis^13^; CD16^+^ classical and IL1B^+^ monocytes in IBD^14-15^; and a shared CXCL10^+^CCL2^+^ inflammatory macrophage population in RA, CD, and UC^16^. Finally, CD90^+^ sublining fibroblasts are expanded in inflammatory arthritis^17-18^. To better understand the causal mechanisms of these disorders, it is crucial to distinguish cell states that genetically regulate autoimmunity from those that represent secondary effects of inflammation.

Autoimmune diseases are highly heritable^19-24^. Consequently, genome-wide association studies (GWAS) have identified thousands of risk loci for autoimmunity^25-30^. However, most of the heritability (outside of the major histocompatibility complex) implicates weak polygenic effects that have not yet been attributed to specific loci^31^. In autoimmunity, heritability is enriched in regulatory regions and around gene bodies of T-cell-specific genes^32-37^. Therefore, it is likely that many loci not reaching genome-wide significance have subtle effects on pathogenic immune cell states^38^. Although methods to partition heritability have pointed to disease-critical cell types^33,39^, they are unable to systematically point to fine-grained cell states and often require making peak-to-gene linking assumptions. A key challenge is that the heritability of autoimmunity has been best captured through open chromatin regions^37,40^, which reflect regulatory activity, whereas recently discovered cell states in tissue inflammation have been defined with transcriptional and surface protein marker data^41-42^.

One as-yet unexplored strategy to capture heritability is to map “dynamic” peaks: chromatin regions whose accessibility changes across transcriptionally defined cell states within a cell type. Disease heritability that can be mapped to these dynamic peaks may identify cell-state-specific gene regulation in disease, and, by doing so, implicate specific cell states. In this work, we used multimodal snRNA-seq and snATAC-seq data to identify dynamic peaks that are associated with transcriptional variation reflecting cell state. We investigated the extent to which dynamic peaks and their associated cell states account for the heritability of 12 autoimmune and allergic diseases, using their genome-wide association study (GWAS) summary statistics (**Fig. 1**).

**Fig. 1.**
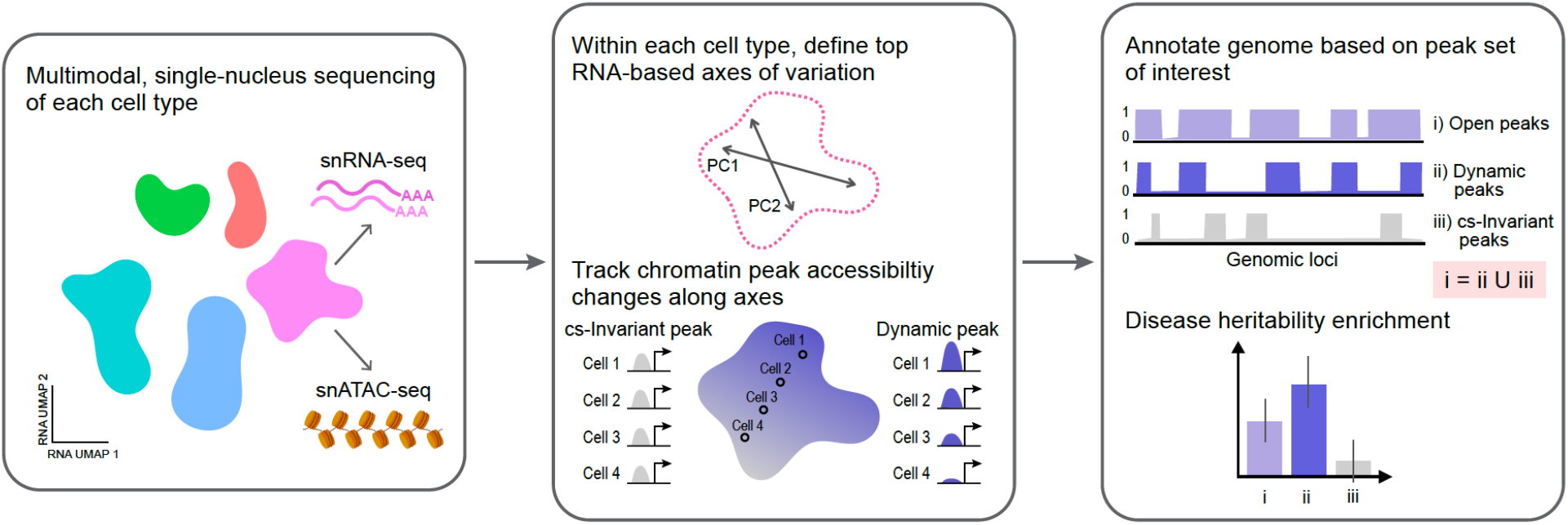
Overview of approach: identification of dynamic regulatory elements from single-nucleus multimodal data marks genomic loci enriched for disease heritability. We collected multimodal single-nucleus RNA-seq and ATAC-seq data from the inflamed synovial tissue of 12 donors (11 with RA, 1 with osteoarthritis) across five cell types (B cells, T cells, endothelial cells, myeloid cells, and fibroblasts), spanning 28,674 cells. Within each of the five cell types, we defined the top snRNA-based axes of variation, e.g., principal components. Then, separately for each cell type, we used a multivariate Poisson generalized linear model to identify the regions of open chromatin whose accessibility were significantly associated with transcriptional variation. We defined associated, cell-state-dependent peaks as “dynamic” (purple) and non-associated peaks as cell-state-invariant, or “cs-invariant” (grey). The purple gradient indicates the accessibility profile for a peak that is associated with both PC1 and PC2. Then, we created SNP-based genomic annotations indicating membership in each peak set (dynamic or cs-invariant). Finally, we partitioned the heritability of 19 polygenic traits (including 12 autoimmune and allergic diseases) and identified peak sets and cell states that were enriched for disease-associated genetic variation.

The hallmark of RA is inflammation in the synovium, a thin membrane surrounding the joint space that expands with immune cell infiltration^4^. We examined cell state data from inflamed synovial tissue as a model for autoimmune diseases at large. We have previously demonstrated that inflammatory cell states in the synovium are seen in tissues of other autoimmune diseases^16-17^. We applied multimodal snRNA and snATAC sequencing to synovial biopsies from 12 donors (11 with RA, 1 with osteoarthritis)^43^. We obtained data on 22,537 genes and 132,466 peaks for 28,674 cells (after quality control), including 7942 T, 1543 B, 7320 myeloid, 9902 fibroblast, and 1967 endothelial cells (**Fig. 2a, Supplementary Fig. 1, Methods**).

**Fig. 2.**
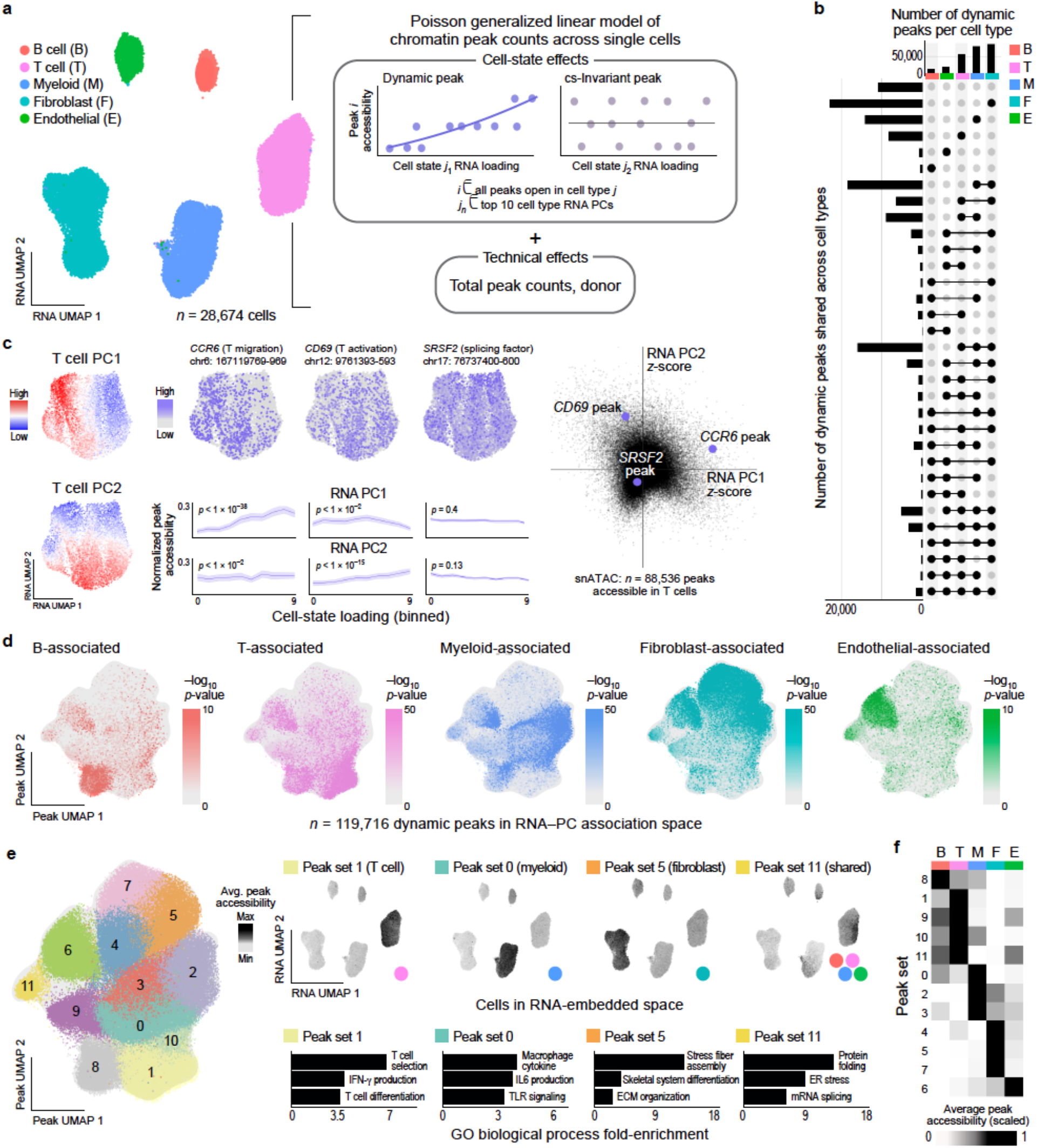
Dynamic peaks implicate cell-type-specific genes and regulatory processes. **a**, (left) snRNA-seq UMAP of all five cell types assayed in this study (n = 28,674 cells), colored by reference-mapped broad cell types (**Methods**). (right) For each cell type, we use a Poisson GLM to identify dynamic peaks whose accessibility is correlated with the top 10 snRNA-seq donor-harmonized principal components (PCs) after accounting for covariates. **b**, Frequency of membership of dynamic peaks to each cell type. **c**, (left) Example of two dynamic peaks respectively associated with the first two RNA PCs in T cells and one cell-state-invariant (“cs-invariant”) peak, as well as (right) the z-scores for PC1 and PC2 of all accessible T cell peaks. Peak windows are 200 base pairs, as noted on the UMAP for each peak. **d**, UMAP of detected dynamic peaks for each cell type, based on a matrix of peaks by z-scores for each of 10 PCs for each of 5 cell types. **e**, (left) UMAP of all dynamic peaks within each cell type colored by Leiden cluster membership, defined by z-scores for each of 10 PCs for each of 5 cell types. (right) For four of these clusters (*i*.*e*., peak sets), we show the snRNA-seq UMAPs of all five cell types shown in panel a, shaded by the mean accessibility of all peaks in the cluster. Color of dot in lower right indicates representative cell type for peak set. Top GO biological processes that are enriched for genes whose promoters (TSS +/- 1 kb) overlap peaks in the four selected clusters (FDR 5%). Supplementary Tables 7-18 include all significant processes for all 12 peak sets. TLR: toll-like receptor; IL-6: interleukin-6. **f**, Heatmap showing the average accessibility of peaks in each peak set scaled by cell type, for each of the 12 peak sets and the five cell types studied.

We used these data to identify cell-state-dependent snATAC-seq peaks. For each cell type, we first defined axes of RNA variation by calculating the top ten principal components (PCs) in transcriptional space, removing batch effects with Harmony (**Supplementary Fig. 2, Methods**)^44^. Corrected RNA PCs are commonly used to define functional states in single-cell analysis. Then, individually for each cell type, we identified cell-state-dependent snATAC-seq peaks as those whose signal can be explained by these ten RNA PC values. We used a multivariable Poisson model, accounting for donor effects and total peak counts (**Fig. 2a, Methods**), to obtain (i) robust likelihood-ratio-test p-values (p_cellType_) to quantify the model’s global significance and (ii) Wald p-values (p_PC_) to quantify the significance of each PC (**Methods**). We tested peaks that were accessible in ≥50 cells in that cell type. Permutation analysis demonstrated that p_cellType_ was not inflated under the null (**Supplementary Fig. 3a, Methods**). We denoted peaks with a global FDR_cellType_<0.05 within each cell type as “dynamic” and all other accessible peaks as cell-state-invariant, or “cs-invariant” (**Fig. 2a, Supplementary Tables 1-5**).

**Fig. 3.**
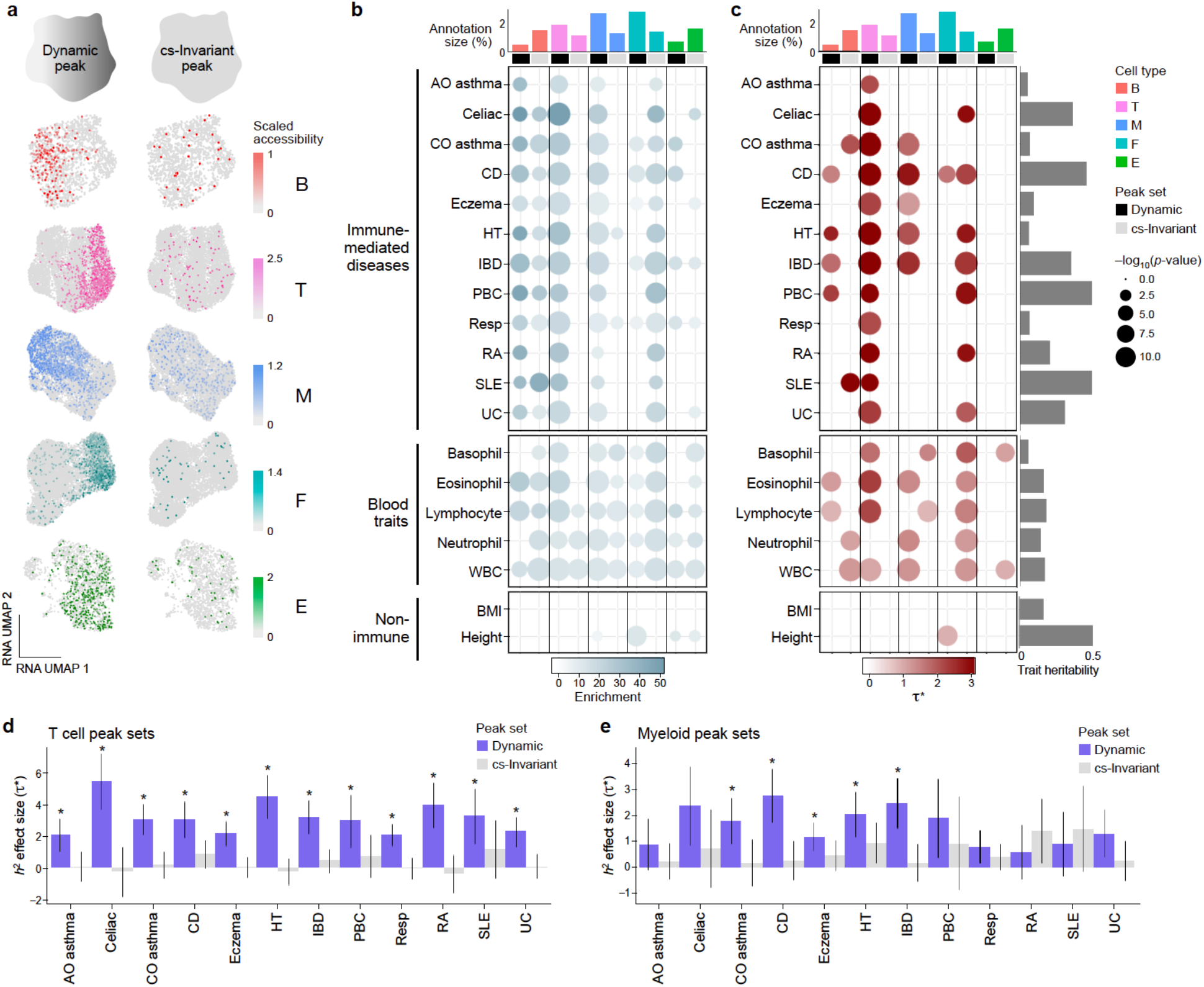
Dynamic peaks are more strongly enriched for disease heritability than cs-invariant peaks. **a**, An example cell-state-dynamic (“dynamic”) and cell-state-invariant (“cs-invariant”) peak for each cell type. Dynamic and cs-invariant peaks plotted are as follows: B (chr8:71843830-71844030 and chr17:40139451-40139651), T (chr2:86786271-86786471 and chr8:11199555-11199755), M (chr11:915102-915302 and chr3:56988979-56989179), F (chr2:217846347-217846547 and chr3:71985293-71985493), and E (chr6:170011289-170011489 and chr1:86793165-86793365). **b**, Heritability enrichment for dynamic (black) and cs-invariant (grey) peaks within each cell type, across 19 complex traits and polygenic diseases. Dot size is proportional to -log_10_(p-value); color intensity is proportional to enrichment. **c**, Estimate of the conditional annotation effect size (*τ*\*) for each peak set; dynamic (black) and cs-invariant (grey) peak annotations are jointly modeled. For panels **b** and **c**, data are plotted only for significant results using a Bonferroni correction threshold of 0.05/19. **d**, Comparison of *τ*\* annotation effect sizes between dynamic (purple) versus cs-invariant (grey) peak-based annotations in T cells. Error bars represent 95% confidence intervals. **e**, Same as in d, but for myeloid cell peak sets. For panels **d** and **e**, asterisks indicate *τ*\* p<0.05/19. Trait acronyms: adult-onset asthma: AO Asthma, child-onset asthma: CO Asthma, Crohn’s disease: CD, hypothyroidism: HT, inflammatory bowel disease: IBD, primary biliary cirrhosis: PBC, respiratory ear-nose-throat disease: Resp, rheumatoid arthritis: RA, systemic lupus erythematosus: SLE, ulcerative colitis: UC.

We observed that 90% (119,716/132,466) of all peaks were dynamic in at least one cell type, and 12% percent of those dynamic peaks (14,365/119,716) were dynamic in all three immune cell types (**Fig. 2b, Supplementary Fig. 3b, Supplementary Tables 1-5**). Both dynamic and cs-invariant peaks lay predominantly in enhancers (111,973/119,716=93.5% and 9,386/10,234=91.7% of peaks, respectively), and dynamic peaks were slightly more common in promoters (defined as 1 kb ± the gene transcriptional start site, TSS, **Methods**) than cs-invariant peaks (odds ratio=1.1, p=0.007). Two dynamic peaks in T cells, chr6:167119769-969 (overlapping T cell migration gene *CCR6*’s promoter) and chr12:9761393-593 (overlapping T cell activation gene *CD69*’s promoter), were significantly associated with a cytotoxicity expression program reflected in PC1 (p_PC1_<1e-38) and memory expression program reflected in PC2 (p_PC2_<1e-15), respectively (**Fig. 2c**). In contrast, a cs-invariant peak (chr17:76737400-600) was near splicing-related gene *SRSF2*’s promoter (**Fig. 2c**). We note that cs-invariant and dynamic peaks had similar read counts (cs-invariant μ=0.096, dynamic μ=0.108 reads) (**Supplementary Fig. 3c**), suggesting our method did not simply classify peaks with sparse signal as cs-invariant.

Next, we investigated whether dynamic peaks could be grouped into functional categories. Since the regression coefficients reflect the strength of association of a peak with a transcriptional component, two peaks with similar coefficients should be active in the same transcriptionally defined cell states. For each peak dynamic in at least one cell type, we obtained its ten RNA PC coefficients for each of the five cell types (n = 50 features) (**Methods**). We visualized variation across these 119,716 peaks with UMAP (**Fig. 2d**) and applied Leiden clustering to define 12 distinct peak sets. Many peak sets included promoters (TSS±1kb) of genes specifically important in one cell type (**Fig. 2e, Supplementary Fig. 4, Methods**). Gene Ontology (GO) analysis revealed that cs-invariant peaks were predominantly enriched for nonspecific processes such as transcription and protein localization (**Supplementary Tables 6, Methods**), whereas most dynamic peak sets were enriched for cell-type-specific processes (**Fig. 2e, Supplementary Tables 7-18, Methods**). Additionally, most dynamic peak sets had increased accessibility in primarily one cell type, further supporting the cell-type-specific nature of dynamic peaks (**Fig. 2f, Supplementary Fig. 4**).

We next hypothesized that dynamic peaks would identify genomic regions capturing more disease heritability than cs-invariant peaks. We examined 12 autoimmune and allergic diseases (average GWAS N = 187,158); as controls, we examined five blood cell traits and two non-immune traits (**Supplementary Table 19, Methods**). We annotated SNPs genome-wide by whether they resided in dynamic and cs-invariant peaks for each cell type (**Fig. 3a, Supplementary Fig. 5a**). We used stratified LD-score regression (S-LDSC)^45^ to partition trait heritability (**Methods**). We used heritability enrichment and standardized annotation effect size (*τ*\*) to evaluate how well our annotations captured causal variation. Enrichment is defined as the proportion of heritability explained by the annotation divided by the number of SNPs in the annotation. The *τ* parameter is proportional to the trait heritability attributed to SNPs in the annotation; *τ*\* corrects for trait heritability and annotation size. As a preliminary check in immune cells, we found that peaks accessible in T cells (both dynamic and cs-invariant) had high *τ*\* across the 12 diseases (p<0.0042=0.05/12, meta-*τ*\*±standard error = 2.5±0.1, individual *τ*\* range=1.9 to 4.6), and those accessible in myeloid and B cells had high *τ*\* values for a subset (**Supplementary Fig. 5b, Supplementary Table 20, Methods**), consistent with previous results^32-34,36-37,39^.

First, we asked whether dynamic peaks had different enrichments than cs-invariant peaks. Meta-analyzing across the 12 immune-mediated diseases, we found that heritability was enriched in dynamic peaks in the three immune cell types (B cell, T cell, and myeloid dynamic meta-enrichment ± SE = 27.7±2.1, 22.0±1.0, and 13.5±0.7, respectively; **Methods**) (**Fig. 3b, Supplementary Table 21**). Autoimmune heritability was also enriched in cs-invariant peaks in B cells (meta-enrichment = 14.9±1.2) and fibroblasts (meta-enrichment = 18.7±1.2) (**Fig. 3b, Supplementary Table 21**). Broadly, dynamic peaks in immune cells tended to capture more heritability and be more enriched for autoimmune disease heritability than cs-invariant peaks. In T and myeloid cells, these results were supported by dynamic peaks capturing between 35-93% and 28-68% of heritability across these 12 diseases, compared to 6-15% and 8-24% of heritability captured by cs-invariant peaks, respectively (**Supplementary Table 21**).

Next, we tested our hypothesis that dynamic peaks capture more autoimmune heritability than cs-invariant peaks. We ran a conditional *τ*\* analysis across the immune-mediated diseases that included both dynamic and cs-invariant peak sets for each cell type (**Methods**). For all three immune cell types, dynamic peaks harbored more heritability than their cs-invariant counterparts (dynamic versus cs-invariant meta-*τ*\*=1.4±0.1 vs 1.3±0.2 for B cells, 2.8±0.2 vs 0.2±0.1 for T cells, and 1.4±0.1 vs 0.5±0.1 for myeloid cells; **Fig. 3c, Supplementary Table 21, Methods**). For T cells, including dynamic peaks in the model obviated any signal in cs-invariant peaks; we observed the largest differences in celiac (dynamic *τ*\*=5.4±0.9 versus cs-invariant *τ*\* -0.2±0.8), HT (4.5±0.7 versus -0.2±0.4), and RA (3.9±0.7 versus -0.4±0.6) (**Fig. 3d, Supplementary Fig. 5c, Supplementary Table 21**). Myeloid and B cells exhibited similar but fewer statistically significant differences between cs-invariant and dynamic peak sets (**Fig. 3e, Supplementary Fig. 5c, Supplementary Table 21**). Since the cs-invariant peak heritability enrichment signal largely disappeared when conditioning on dynamic peaks (**Fig. 3c-e**), we concluded that autoimmune disease heritability in cs-invariant peaks was likely explained by nearby dynamic peaks. That is, dynamic peaks tagged causal variants better than cs-invariant peaks. Additionally, within dynamic peaks in immune cells, we saw high enrichment and *τ*\* values for five blood cell traits (positive controls) and low values for two non-immune traits (negative controls); we note that cs-invariant peaks in fibroblasts retained a significant *τ*\* for height (**Fig. 3c, Supplementary Table 21**).

Our findings suggest that, within T and myeloid cells, specific cell states might drive autoimmunity. Examining the subsets of dynamic peaks that are most active in subpopulations of these broad cell types may distinguish cell states integral to genetic causality from those that are consequences of inflammation. To identify fine-grained, discrete biological cell states within T and myeloid cells, we used Symphony^46^ to map the multimodal snRNA-seq profiles onto established AMP-RA/SLE atlases of 94,048 T and 76,181 myeloid cells, respectively, obtained from RA synovial tissues^49^ (**Methods**). Next, to define the subset of peaks most active within each discrete cell state, we identified the top 25% of dynamic peaks whose regression z-scores most closely resembled the average cell state (centroid) loadings for the top ten within-cell-type RNA PCs (**Supplementary Fig. 6a, Methods**), recognizing that multiple cell states may be linked to the same peak (**Supplementary Fig. 6b, 7**). We then used these peaks to define a cell-state-specific annotation.

For each of the 23 discrete T cell states in our dataset, we applied S-LDSC to estimate the enrichment of disease heritability in dynamic peaks (**Fig. 4a, Methods**). We found that six cell states disproportionately captured RA heritability: CD4^+^ Tfh/Tph (T-3) *τ*\*=3.9±0.9, CD4^+^ Tph (T-7) *τ*\*=3.7±0.9, CD4^+^CD25^hi^ regulatory T (Treg, T-8) *τ*\*=3.0±1.2, CD4^+^CD25^lo^ Treg (T-9) *τ*\*=3.4±1.0, CD4^+^OX40^+^NR3C1^+^ (T-10) *τ*\*=4.4±1.0, and CD4^+^CD145^+^ memory (T-11) *τ*\*=3.8±1.0 (**Fig. 4b, Supplementary Table 22**). We observed a similar pattern across the remaining 11 immune-mediated diseases, with T-7 and T-8 standing out as significant (p<0.05/12) across 10/12 and 11/12 diseases, respectively (**Fig. 4c, Supplementary Tables 22-23**). Meta-analyzing these diseases jointly (**Methods**) resulted in meta-*τ*\* values for T-3=2.0±0.2, T-7=2.0±0.1, T-8=1.9±0.2, T-9=2.0±0.2, T-10=1.9±0.2, and T-11=2.1±0.2 (**Fig. 4d, Supplementary Table 23**). Interestingly, while both Tph (T-3 and T-7) and CD8^+^GZMK^+^ memory T cells (T-13 and T-14) have been shown to be expanded in inflammatory tissues^4,10^, only the former harbored global heritability signal, suggesting that, outside of gut-related diseases, the expansion of the CD8^+^GZMK^+^ population might be a secondary effect of inflammation.

**Fig. 4.**
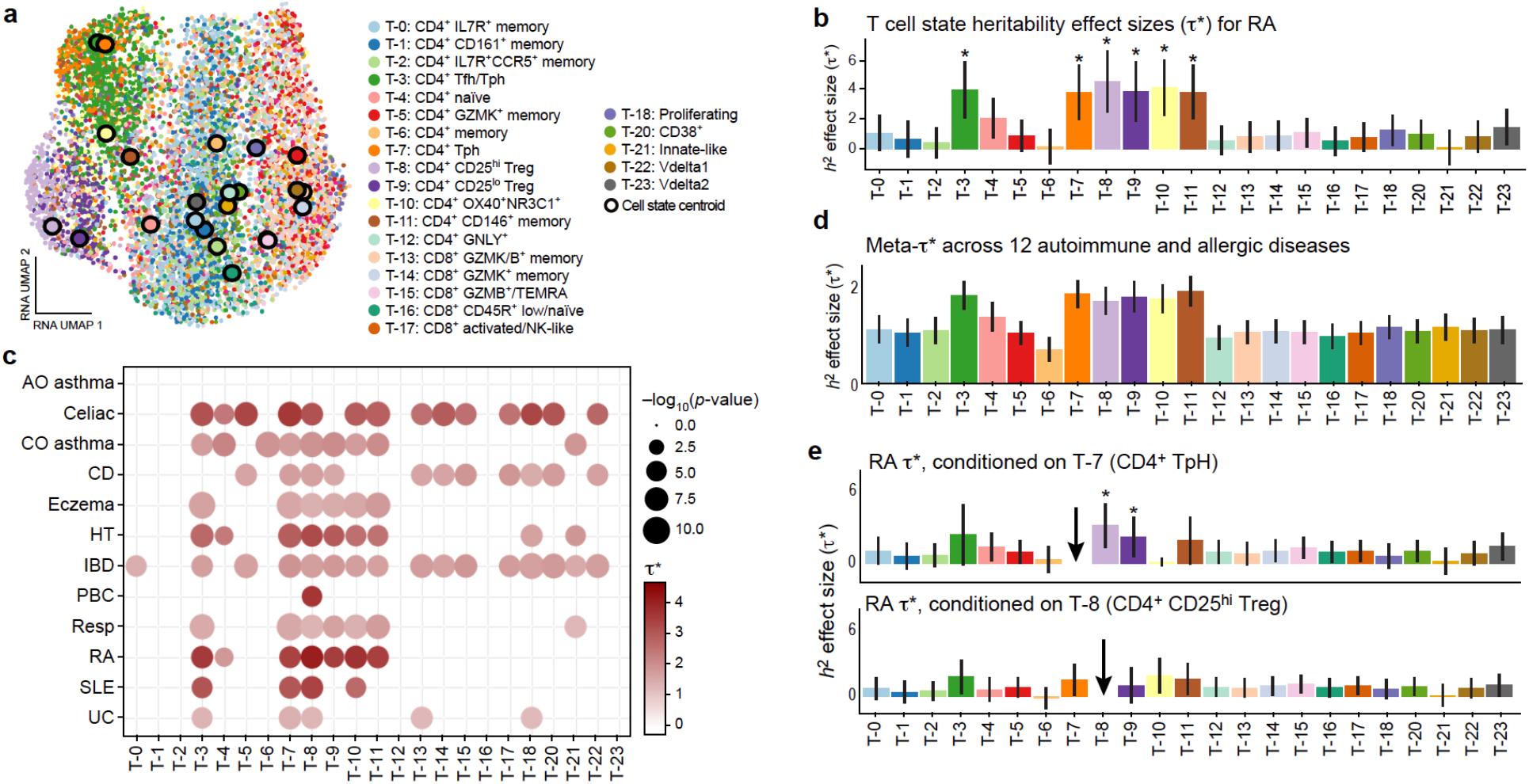
Dynamic peaks help distinguish disease-critical T cell states for immune-mediated disease. **a**, snRNA-seq UMAP of T cells, colored by reference-mapped T cell states (**Methods**). Larger circled dots indicate cell state centroids. **b**, Annotation effect size (*τ*\*) for each discrete T cell state, for rheumatoid arthritis (RA). Error bars indicate +/- 95% CIs, and asterisks indicate p < 0.05/12. **c**, *τ*\* for each T cell state across 12 autoimmune and allergic diseases. Dot size is proportional to -log_10_(p-value), color intensity is proportional to *τ*\*, and only *τ*\* values significant at a Bonferroni correction threshold (p < 0.05/12) are shown. All annotations were defined by the top 25% of dynamic peaks that had the highest cosine similarity to the cell state’s average PC loadings (**Methods**). **d**, Meta-analyzed *τ*\* for each T cell state across all 12 autoimmune and allergic diseases. Error bars indicate +/- 95% CIs. All p-values are <1e-15. **e**, RA *τ*\* for all T cell states as defined in **a** but conditional on either (top) T-7 or (bottom) T-8, the two strongest effect cell states. Error bars indicate +/- 95% CIs, and asterisks indicate p < 0.05/12. Trait acronyms: adult-onset asthma: AO Asthma, child-onset asthma: CO Asthma, Crohn’s disease: CD, hypothyroidism: HT, inflammatory bowel disease: IBD, primary biliary cirrhosis: PBC, respiratory ear-nose-throat disease: Resp, rheumatoid arthritis: RA, systemic lupus erythematosus: SLE, ulcerative colitis: UC.

We then wanted to see what signal might be shared versus unique across the highest-scoring states, since peaks between certain cell states overlapped (**Supplementary Fig. 6b, 7**). Conditioning on the most significant cell state overall, T-7 (meta-*τ*\* p<1e-39), removed signal in T-3, T-10, and T-11 but retained signal in the two Treg populations in RA (**Fig. 4e, Supplementary Fig. 8a, Supplementary Tables 23-24**) and reduced signal for T-3 and T-8 through T-11 in the meta-analysis (**Supplementary Fig. 8b**). Conditioning on the most significant Treg, T-8 (meta-*τ*\* p<1e-33), removed signal in T-3, T-7, and T-9 through T-11 in RA (**Fig. 4e, Supplementary Fig. 8c, Supplementary Tables 23-24**); this signal remained low when meta-analyzing all 12 diseases and significant at p<0.05/12 only for a subset (**Supplementary Fig. 8c-d**). We note that, using a more stringent set of associated peaks (10% versus 25%), the Tfh/Tph and Tph signals were retained when conditioning on either Treg population, and vice versa (**Supplementary Fig. 9a-b**). Overall, these results complement previous studies demonstrating the role of these populations in autoimmunity^4-9,16,47-48^ that highlight Tph and Treg cells as key cell states harboring heritability across many autoimmune conditions.

We similarly examined myeloid cells, identifying 14 discrete myeloid biological states in our data and applying S-LDSC to peak-based annotations (**Fig. 5a, Methods**). We found that in IBD, six myeloid cell states explained a disproportionate amount of heritability: STAT1^+^CXCL10^+^ (M-6) *τ*\*=2.5±0.5, DC3 (M-9) = 2.0±0.5, DC2 (M-10) = 2.5±0.5, DC1 (M-12) = 2.4±0.5, DC4 (M-11) = 2.2±0.5, and LAMP3^+^ (M-14) = 2.9±0.5 (**Fig. 5b, Supplementary Table 25**). For these states, we observed similar patterns of enrichment across the 11 remaining diseases (M-6 meta-*τ*\*=1.9±0.2, M-12=1.7±0.2, M-10=1.8±0.1, M-9=1.3±0.1, M-11=1.3±0.1, and M-14=2.1±0.2), with M-6 and M-14 having p<0.05/12 for 8/12 and 10/12 diseases, respectively, and 3/4 DC populations having p<0.05/12 for the gut-related diseases (Celiac, CD, IBD, and UC) (**Fig. 5c-d, Supplementary Tables 25-26**).

**Fig. 5.**
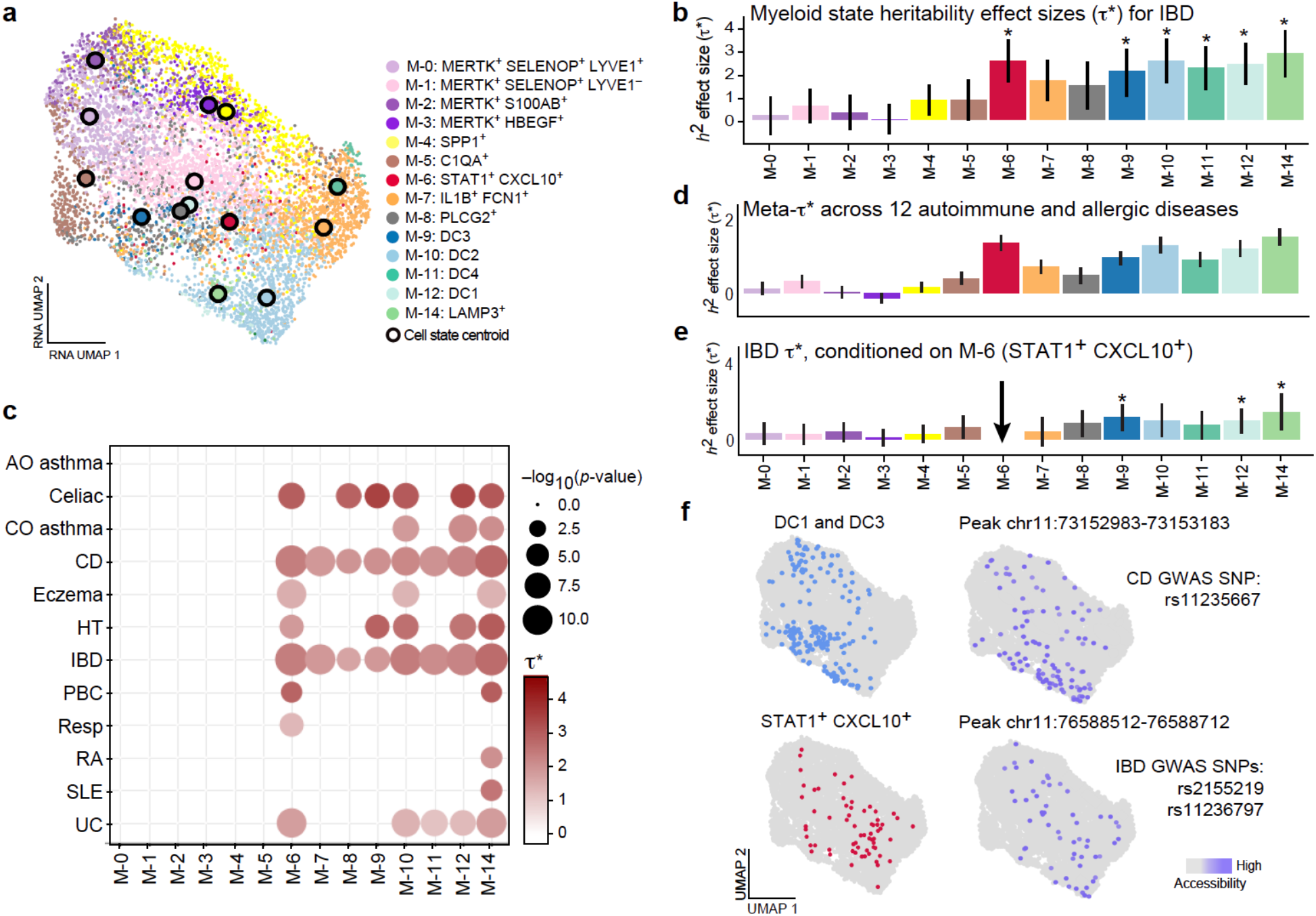
Dynamic peaks help distinguish disease-critical myeloid cell states for immune-mediated disease. **a**, snRNA-seq UMAP of myeloid cells, colored by reference-mapped myeloid cell states (**Methods**). Larger circled dots indicate cell state centroids. **b**, Annotation effect size (*τ*\*) for each discrete myeloid cell state, for inflammatory bowel disease (IBD). Error bars indicate +/- 95% CIs, and asterisks indicate p < 0.05/12. **c**, *τ*\* for each discrete myeloid cell state across 12 autoimmune and allergic diseases. Dot size is proportional to -log_10_(p-value), color intensity is proportional to *τ*\*, and only *τ*\* values significant at a Bonferroni correction threshold (p < 0.05/12) are included. All annotations were defined by the top 25% of dynamic peaks that had the highest cosine similarity to the cell state’s average PC loadings (**Methods**). **d**, Meta-analyzed *τ*\* for each myeloid cell state across 12 autoimmune diseases. Error bars indicate +/- 95% Cis. All p-values for M-6 through M-14 are <1e-5. **e**, *τ*\* for IBD, as in panel **a**, but conditional on M-6 cells. Asterisks indicate p<0.05/12. **f**, Two examples of (Left) high-scoring cell states and (Right) an example associated peak’s accessibility profile for each cell state. Of note, SNPs in both peaks have been linked to autoimmune diseases through previous IBD and CD GWAS studies (SNP rsID listed on the right).

Notably, M-6, previously shown to be expanded in inflammation^16^, was one of the most significant states across all 12 diseases (meta-*τ*\* p<1e-38). Conditioning on M-6 obviated signal for most other populations in IBD, retaining signal only in M-9, M-12, and M-14, the latter of which shares 71% (13,724/19,232) of its peaks with M-6 (**Fig. 5e, Supplementary Fig. 6c**,**10**). In the meta-analysis, conditioning on M-6 removed signal from all other states across diseases, although a few individual traits such as CD and HT retained p<0.05/12 for M-9 and M-12 (**Supplementary Fig 8e-f, Supplementary Tables 24**,**26**). Using the top 10% of peaks only retained signal in M-9 and M-12 (**Supplementary Fig. 9c-d**), indicating some potentially distinct heritability captured by these DC populations.

Finally, for a few traits and myeloid cell states with high *τ*\* values, we assessed whether SNPs previously associated with autoimmune conditions were also associated with these states. One example is the SNP rs11235667 (chr11:73152652), which has been associated in a GWAS for CD^50^. This risk locus also falls within a peak that is in the top 10% of similar peaks for both M-9 and M-12 (chr11:73152483-73153683), the two DC populations that retained signal after conditioning on M-6 (**Fig. 5f**). Additionally, two genome-wide risk loci for IBD—rs2155219 (chr11:76588150)^51^ and rs11236797 (chr11:76588605)^52^—fall within peak chr11:76588012-76589212, which is in the top 10% of similar peaks for M-6 (**Fig. 5f**). Our results suggest that the highlighted cell states may mediate genetic risk for autoimmunity via their associated peaks.

Here, we present a novel paradigm in which we identified and leveraged dynamic regulatory elements within cell types to capture disease heritability in single-cell data. Our work provides a principled approach to linking cell states observed in a tissue context with heritability signals. This reflects an advance over previous heritability enrichment analyses–which have largely implicated cell types but have lacked the resolution to implicate cell states. While we used cells sampled from the inflamed RA synovium, analyzing multimodal data from multiple inflammatory diseases could be even more informative, since they may capture an even broader spectrum of tissue-resident immune cell states. This approach has the potential to be applied to other traits where multimodal snRNA-seq and snATAC-seq data can be derived from critical tissues. Identification of causal cell states using heritability enrichment could be valuable broadly across complex traits for developing therapies that target specific, causal cell populations. Our framework will become increasingly powerful as both tissue-based, single-cell multimodal and GWAS studies increase in scale and scope.

## Methods

### Sample collection, multimodal sequencing, and counts matrices preparation

The RA tissue 10x Genomics Single Cell Multiome ATAC + Gene Expression datasets used in this study are described in detail in Weinand et al.^43^. Briefly, we collected synovial tissues from 11 Rheumatoid Arthritis patients and 1 Osteoarthritis patient (n = 12 donors).These samples were cryopreserved, thawed, and disaggregated into single-cell suspensions whose nuclei were subsequently isolated as previously described^53-54^.

Joint scRNA-seq and scATAC-seq libraries were prepared using the 10x Genomics Single Cell Multiome ATAC + Gene Expression kit according to manufacturer’s instructions. Each sample was processed individually. Libraries were sequenced on an Illumina Novaseq to a target depth of 20,000 read pairs per nuclei for mRNA libraries and 25,000 read pairs per nucleus for the ATAC libraries. Initial processing was done in Cell Ranger ARC pipeline (version 2.0.0) from 10x Genomics. To deduplicate ATAC reads from PCR amplification bias within a cell while keeping reads originating from the same positions but from different cells, we used in-house scripts^43^.

We applied stringent quality control (QC) measures to both the nuclear RNA and ATAC data^43^. Briefly, RNA cells had to pass Cell Ranger ARC cell calling, >500 genes, and <20% mitochondrial reads. For ATAC features, we used an in-house well-curated consensus RA tissue peak set from a larger study of RA synovial tissue scATAC-seq data^43^. ATAC cells had to have at least 10,000 post read-QC reads, with >50% of reads falling in peak neighborhoods, >10% in promoters (TSS-2kb), <10% in mitochondrial chromosome, and <10% in ENCODE blacklist^55^. Doublets were removed within modalities^43^. We thus obtained count matrices of gene expression and ATAC peaks with corresponding cell barcodes.

### Cell type labeling and data preprocessing

To assign broad cell type labels, we used canonical correlation analysis (CCA) between the cells by genes and cells by peaks matrices; each matrix was normalized (log(CP10K) and log(TFxIDF), respectively), had most variable features selected, and scaled before running. The resulting cells by peak canonical variate matrix was batch corrected by sample using Harmony^44^ and Louvain clustered. Cell types were determined using marker peaks falling within the promoters of the following cell type marker genes: *CD3D* in T cells, *MS4A1* in B cells, *C1QA* in myeloid cells, *PDPN* in fibroblasts, and *VWF* in endothelial cells. If multiple peaks overlapped a gene’s promoter, the peak whose accessibility best tracked with the genes’ expression was used. Within each broad cell type, we removed any cells in which the corresponding donor had less than 5 cells.

For each broad cell type, we used Symphony^46^ to map the multiome snRNA-seq cells into the harmonized reference space generated from the AMP-RA synovial tissue CITE-seq cells^49^. We chose the most common cell state for each multiome cell’s 5 nearest neighbors among the AMP-RA CITE-seq cells. We excluded any multiome T cells that mapped to the cell state that was defined as having a high number of mitochondrial reads (“MT-high”). We also excluded the 4 cells that mapped to the myeloid cell state plasmacytoid dendritic cells (M-13). These steps yielded the following numbers of cells per cell type: T (7,942), B (1,543), myeloid (7,320), fibroblast (9,902), endothelial (1,967).

For the snATAC-seq data used for regressions, we excluded peaks that were accessible in <50 cells in each cell type and otherwise left the counts as-is. All remaining peaks were included in our “OPEN” peak set annotation for each cell type. For snATAC-seq data used to track normalized peak accessibility (as in **Figure 2e-f**), we binarized the peaks x cells matrix, ran log(TFxIDF) normalization, selected the most variable peaks, and centered and scaled features to mean 0 and variance 1 across cells.

For the snRNA-seq data, which we used to determine the RNA PCs (*i*.*e*., cell-state axes of variation), we excluded genes expressed in <50 cells of a given cell type and ribosomal and mitochondrial genes from our analyses. To account for differences in library size and read depth, we then normalized counts such that the total number of transcripts per cell was scaled to counts per 10,000 (CP10K). We took the log1p of these scaled counts and, keeping only the top 5,000 most variable genes in each cell type, variance scaled genes such that they were zero centered and had unit variance. We calculated the top 20 PCs in the RNA space, upon which we ran Harmony^44^ to correct for donor-specific effects. We used the donor-harmonized RNA PCs for all downstream analyses, including all regressions and UMAP visualizations.

### Identifying dynamic, cs-invariant, and PC-associated peaks

After computing the principal components from snRNA-seq, we used a multivariable Poisson regression model to predict each snATAC-seq peak’s accessibility as a function of the top ten snRNA-seq principal components and technical covariates while accounting for the sparsity and asymmetry of the snATAC-seq peak counts:

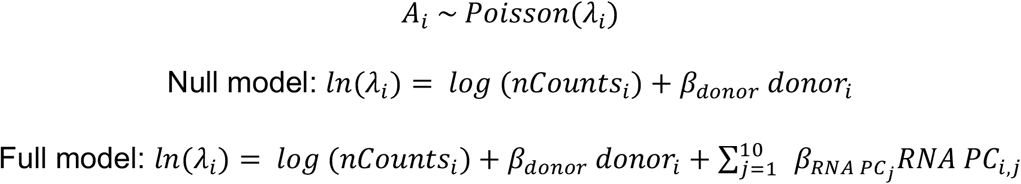

where *A* is the peak’s accessibility (read counts) in cell *i, nCounts* is the total counts in cell *i*, and *donor* is the donor ID of cell *i*, one-hot encoded into 12 variables. We model the total counts as an offset and donor membership as a fixed effect. *RNA PC*_*i,j*_ is the PC value for cell *i* with respect to *RNA PC*_*j*_, where *j* ranges from 1 to 10. We ran this regression for every accessible peak in each of the five cell types studied. We identified ATAC peaks whose accessibility is associated with these expression PCs, determining significance based on the difference between the full and null models at a 10-degree-of-freedom likelihood ratio test (LRT) at 5% FDR within each cell type. We called peaks with FDR < 0.05 “dynamic”; those peaks that were accessible but had FDR ≥ 0.05 were classified as “cs-invariant”. Additionally, we identified peaks that were significantly associated with individual PCs as those that were 1) dynamic based on the overall joint regression and 2) had a nominal Wald p<0.05 for the individual PC in question.

To test for inflated statistics that may have arisen from technical structure, we ran permutation-based checks. Specifically, we shuffled the cell-PC values across all cells within each donor and re-ran the Poisson GLM for every peak to derive permutation-based p-values. We compared the p-value distributions from the permuted versus real models in Q-Q plots (for each of the top 10 PCs in each of the five cell types) against the null (expected, uniform) p-value distribution and calculated inflation using the function at slowkow.com/notes/ggplot2-qqplot.

### Characterizing dynamic peaks

We defined dynamic peaks for each cell type as those with LRT FDR < 0.05 from the joint Poisson GLM (as described above). For each dynamic peak, we recorded the PC-specific coefficient z-score (normalized β) and p-value (p_PC_). To identify potential biological functions, we also took a set of canonical marker genes known to have functional roles in T cells^41^ and identified peaks that overlapped these genes’ promoter regions (transcriptional start site +/- 1 kb), which enabled us to connect specific dynamic peaks with putative biological roles.

To further characterize the cell-type-specificity and potential biological processes represented by peak sets, we defined 50-feature vectors for each peak. Specifically, we combined the ten regression z-scores (one per RNA PC) for each of the five cell types for each peak. For cell types where the peak was not accessible, we assigned z-scores of zero. We calculated UMAP coordinates (where each dot is a peak) using these 50 values for each peak, which allowed visualizing cell-type-significance on a per-peak level.

We then ran Leiden clustering (resolution=1, most broad) on this 50-feature vector X ∼130,000 peaks matrix. For each of the Leiden clusters (which we call a “peak set”), we calculated the mean peak accessibility in each cell and plotted this on the all-cell-type UMAP to assess any cell type differences in overall peak set accessibility. We also identified the set of genes whose promoters overlapped the peaks in each peak set and ran these gene sets through Gene Ontology (geneontology.org) to look for biological processes that were significantly enriched (FDR<0.05) in these peak sets. Finally, we calculated the mean peak accessibility within each peak set in each cell type, scaling across cell types to get values that ranged from 0 to 1.

### Partitioning heritability for each within-cell-type annotation with S-LDSC

We analyzed 19 traits total (average N=280,785) from a subset of 188 traits with publicly accessible summary statistics: 12 autoimmune diseases, six blood-related traits, and two non-immune traits (**Supplementary Table 19**). We chose these specifically given our focus on immune-mediated diseases; blood traits serve as a positive control given our use of many immune cells, and non-immune traits serve as a negative control, given we don’t expect them to involve most of the cell types in our study. Additionally, we removed traits with Z<4.5 (as computed by S-LDSC with baseline LD v2.2), to ensure robustness of our results, and for traits with multiple studies present, we selected the study with the higher N (number of affected individuals) and M (number of individuals overall), which tended to be more recent. The full set of characteristics describing the studies for the traits we analyzed, including references to where they were originally published, are included in **Supplementary Table 19**.

We apply S-LDSC (stratified linkage disequilibrium score regression) (version 1.0.0), a method developed to partition polygenic trait heritability by one or more functional annotations, to quantify the contribution of our defined regulatory annotations to the 19 complex traits mentioned above. We annotate common SNPs (MAF ≥ 0.05) based on peak sets of interest (i.e., dynamic peaks in T cells). Specifically, we binarize the set of peaks with FDR<0.05 (for dynamic peaks) and expand the 200bp peak window by 500 bp on either side to result in a 1.2kb genomic window for each peak. Then, we apply S-LDSC once to the annotated SNPs (converting from hg38 to hg19, to match the summary statistics genome version) to compute population-specific LD scores and again to quantify the complex trait heritability captured by our peak-based annotations. Here, the two statistics we use to evaluate how well our annotations capture causal variation are enrichment and standardized effect size (*τ*\*), as previously defined^32,45^.

Briefly, we calculate enrichment as follows:

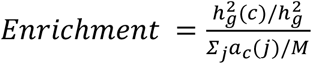

where *h*^*2*^_*g*_*(c)* is the heritability explained by SNPs in annotation *c, h*^*2*^_*g*_ is the overall heritability of a trait, *a*_*c*_ is the value of annotation *c, j* is SNP *j*, and *M* is the number of common (MAF≥0.05) SNPs (5,961,159 in Europeans and 5,469,053 in East Asians). Whereas enrichment does not quantify effects that are unique to a given annotation, *τ*\* does (i.e. if conditioning two correlated annotations in a joint S-LDSC model, they will have similar enrichments, but the *τ*\* for the annotation with greater true causal variant membership will be larger and more significantly positive). We calculate *τ*\* as follows:

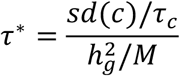

where *sd(c)* is the standard deviation of the annotation *c, τ*_*c*_ is the per-SNP contribution of one unit of the annotation *c* to heritability, and *h*^*2*^_*g*_ and *M* are the same as before. Since *τ* is not comparable across annotations or traits, we use *τ*\*, which is comparable and is defined as the per-annotation standardized effect size, or the proportionate change in per-SNP *h*^*2*^ associated with a one standard deviation increase in the value of the annotation.

Each S-LDSC analysis conditions our peak set annotations on 69 baseline annotations, a subset of the 75 annotations referred to as the baseline-LD model as done previously^32^; we removed six annotations including T cell enhancers. The 69 annotations consist of 53 cell-type-nonspecific annotations, 7 which include histone marks and open chromatin, 10 MAF bins, and six LD-related annotations to assess whether functional enrichment is cell-type-specific and to control for the effect of MAF and LD architecture. Consistent inclusion of MAF and LD-associated annotations in the baseline model is the standard recommended practice of using S-LDSC.

For dynamic versus cs-invariant peak sets comparisons, we ran a conditional *τ*\* enrichment, including both dynamic and cs-invariant annotations in a joint run of S-LDSC. We report error bars that reflect 95% confidence intervals around the estimated *τ*\* values and show in heatmaps only those values that remain significant upon Bonferroni correction for multiple hypothesis testing across the 19 traits analyzed.

For the meta-enrichment and meta-*τ*\* calculations across multiple traits for a given annotation, we used an inverse variance weighted analysis, aggregating either individual enrichment values or *τ*\* values, respectively. We ran the R package rmeta (version 3.0, https://github.com/cran/rmeta/blob/master/R/meta.R) and reported the meta summaries estimate and standard error values. Given a sequence of independent observations y_i_ with variances σi^2^, the (i) inverse-variance weighted average and (ii) its variance are given by:

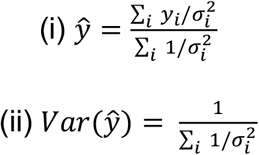

### Building cell state annotations in T and myeloid cells

We defined an annotation for each cell state based on the set of peaks most similar in their accessibility profiles as the cell state. Each peak has an effect size *β*_*j*_ for each RNA PC *j*. Given an individual cell’s RNA-seq profile, we scored each peak for its relevance by calculating the cosine similarity between that peak’s PC z-scores (normalized *β*s) and the cell’s values across the 10 PCs:

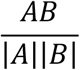

Where *A =* [z-score_RNA PC1_ … z-score_RNA PC10_] for a given peak and *B* = [PC value_RNA PC1_ … PC value_RNA PC10_] for a given cell. This approach can be applied to an individual cell or the average of a collection of cells that form a cluster representing a specific cell state; we chose the latter for computational efficiency. Once scored, we built a cell-state-specific annotation, by identifying the top 25% of peaks with the highest score for each cell state. This allowed for equally sized annotations across cell states within a cell type.

### Identifying previously implicated autoimmune GWAS SNPs

We used the GWAS catalog (https://www.ebi.ac.uk/gwas/) to identify SNPs that had known genetic associations to autoimmune diseases (genome-wide p<5e-8). We then checked for overlap between these SNPs and cell-state-defining peaks and report a few examples as case studies.

## Data Availability

This manuscript uses multimodal single-nucleus data that will be released upon publication of the following manuscript: Weinand et al. (in preparation).

## Code Availability

This work uses the S-LDSC software (https://github.com/bulik/ldsc) to process GWAS summary statistics. All code related to processing of the multimodal single-nucleus data, post-hoc heritability analyses, and generation of the tables and figures in this manuscript are available on Github (https://github.com/gupta-anika/dynamic-heritability).

## Acknowledgments

We thank Kushal Dey, Karthik Jagadeesh, Martin Zhang, Joyce Kang, Samuel Kim, Benjamin Strober, Po-Ru Loh, Shamil Sunyaev, and Laurie Rumker for helpful conversations and Leslie Gaffney for help with figures. This work is supported in part by funding from the National Institutes of Health (R01AR063759, U01HG012009, UC2AR081023, P01AI148102, U19AI095219, R01AR063709, T32AR007530, T32AR007530) and the Broad Institute. K. Wei is supported by a Burroughs Wellcome Fund Career Award for Medical Scientists and a Doris Duke Foundation Clinical Scientist Development Award.

## Author Contributions

A.G. and S.R. conceptualized the study. A.G. conducted all analyses on genetic and post-QC multimodal data. S.R. and T.A. supervised the study. K. Weinand conducted multimodal data quality control and data processing. T.A., A.N., A.L.P., S.S., and AMP RA/SLE provided input on statistical analyses and study design. L.D. and K. Wei recruited patients for multimodal samples, and K. Wei supervised sample processing and single-cell multimodal data generation. A.G., T.A., and S.R. wrote the initial manuscript. All authors contributed to editing the final manuscript.

## References

1. Szekanecz, Z. et al. Autoinflammation and autoimmunity across rheumatic and musculoskeletal diseases. Nature Reviews Rheumatology 17, 585–595 (2021).

2. Rosenblum, M. D., Remedios, K. A. & Abbas, A. K. Mechanisms of human autoimmunity. Journal of Clinical Investigation 125, 2228–2233 (2015).

3. Wang, L., Wang, F.-S. & Gershwin, M. E. Human autoimmune diseases: A comprehensive update. Journal of Internal Medicine 278, 369–395 (2015).

4. Rao, D. A. et al. Pathologically expanded peripheral T helper cell subset drives B cells in rheumatoid arthritis. Nature 542, 110–114 (2017).

5. Zhang, F. et al. Defining inflammatory cell states in rheumatoid arthritis joint synovial tissues by integrating single-cell transcriptomics and mass cytometry. Nature Immunology 20, 928–942 (2019).

6. Argyriou, A. et al. Single cell sequencing identifies clonally expanded synovial CD4+ Tph cells expressing GPR56 in rheumatoid arthritis. Nature Communications 13, 4046 (2022).

7. Uzzan, M. et al. Ulcerative colitis is characterized by a plasmablast-skewed humoral response associated with disease activity. Nature Medicine 28, 766–779 (2022).

8. Christophersen, A. et al. Distinct phenotype of CD4+ T cells driving celiac disease identified in multiple autoimmune conditions. Nature Medicine 25, 734–737 (2019).

9. Akama-Garren, E. H. et al. Follicular T cells are clonally and transcriptionally distinct in B cell-driven mouse autoimmune disease. Nature Communications 1, 12 (2021).

10. Jonsson, A. H. et al. Granzyme K+ CD8 T cells form a core population in inflamed human tissue. Science Translational Medicine 14, 649 (2022).

11. Jaeger, N. et al. Single-cell analyses of Crohn’s disease tissues reveal intestinal intraepithelial T cells heterogeneity and altered subset distributions. Nature Communications 12, 1921 (2021).

12. Arazi, A. et al. The immune cell landscape in kidneys of patients with lupus nephritis. Nature Immunology 20, 902–914 (2019).

13. Nakamizo, S. et al. Single-cell analysis of human skin identifies CD14+ Type 3 dendritic cells co-producing IL1B and IL23A in psoriasis. Journal of Experimental Medicine 9, 218 (2021).

14. Liu, H. et al. Subsets of mononuclear phagocytes are enriched in the inflamed colons of patients with IBD. BMC Immunology 1, 20 (2019).

15. Mitsialis, V. et al. Single-cell analyses of colon and blood reveal distinct immune cell signatures of ulcerative colitis and Crohn’s disease. Gastroenterology 159, 591–608 (2020).

16. Zhang, F. et al. IFN-γ and TNF-α drive a CXCL10+ CCL2+ macrophage phenotype expanded in severe COVID-19 lungs and inflammatory diseases with tissue inflammation. Genome Medicine 13, 64 (2021).

17. Wei, K. et al. Notch signaling drives synovial fibroblast identity and arthritis pathology. Nature 582, 259–264 (2020).

18. Zhang, F. et al. Defining inflammatory cell states in rheumatoid arthritis joint synovial tissues by integrating single-cell transcriptomics and mass cytometry. Nature Immunology 20, 928–942 (2019).

19. Seldin, M. F. The genetics of human autoimmune disease: A perspective on progress in the field and future directions. Journal of Autoimmunity 64, 1–12 (2015).

20. Stahl, E. A. et al. Bayesian inference analyses of the polygenic architecture of rheumatoid arthritis. Nature Genetics 44, 483–489 (2012).

21. Redondo, M. J., Jeffrey, J., Fain, P. R., Eisenbarth, G. S. & Orban, T. Concordance for islet autoimmunity among monozygotic twins. New England Journal of Medicine 359, 2849–2850 (2008).

22. Guerra, S. G., Vyse, T. J. & Cunninghame Graham, D. S. The genetics of lupus: A functional perspective. Arthritis Research & Therapy 14, 211 (2012).

23. Gordon, H., Trier Moller, F., Andersen, V. & Harbord, M. Heritability in inflammatory bowel disease. Inflammatory Bowel Diseases 1, 1428–1434 (2015).

24. Selmi, C. et al. Primary biliary cirrhosis in monozygotic and dizygotic twins: Genetics, epigenetics, and environment. Gastroenterology 127, 485–492 (2004).

25. Okada, Y. et al. Genetics of rheumatoid arthritis contributes to biology and drug discovery. Nature 506, 376–381 (2013).

26. Raychaudhuri, S. et al. Five amino acids in three HLA proteins explain most of the association between MHC and seropositive rheumatoid arthritis. Nature Genetics 44, 291–296 (2012).

27. Wang, Y.-F. et al. Identification of 38 novel loci for systemic lupus erythematosus and genetic heterogeneity between ancestral groups. Nature Communications 12, 772 (2021).

28. Patsopoulos, N. A. et al. Multiple sclerosis genomic map implicates peripheral immune cells and microglia in susceptibility. Science 365, 6460 (2019).

29. Liu, J. Z. et al. Association analyses identify 38 susceptibility loci for inflammatory bowel disease and highlight shared genetic risk across populations. Nature Genetics 47, 979–986 (2015).

30. de Lange, K. M. et al. Genome-wide association study implicates immune activation of multiple integrin genes in inflammatory bowel disease. Nature Genetics 49, 256–261 (2017).

31. Manolio, T. A. et al. Finding the missing heritability of complex diseases. Nature 461, 747–753 (2009).

32. Amariuta, T. et al. Impact: Genomic annotation of cell-state-specific regulatory elements inferred from the epigenome of bound transcription factors. The American Journal of Human Genetics 104, 879–895 (2019).

33. Finucane, H. K. et al. Heritability enrichment of specifically expressed genes identifies disease-relevant tissues and cell types. Nature Genetics 50, 621–629 (2018).

34. Hu, X. et al. Integrating autoimmune risk loci with gene-expression data identifies specific pathogenic immune cell subsets. The American Journal of Human Genetics 89, 496–506 (2011).

35. Farh, K. K.-H. et al. Genetic and epigenetic fine mapping of causal autoimmune disease variants. Nature 518, 337–343 (2014).

36. Trynka, G. et al. Chromatin marks identify critical cell types for fine mapping complex trait variants. Nature Genetics 45, 124–130 (2012).

37. Gusev, A. et al. Partitioning heritability of regulatory and cell-type-specific variants across 11 common diseases. The American Journal of Human Genetics 95, 535–552 (2014).

38. Boyle, E. A., Li, Y. I. & Pritchard, J. K. An expanded view of complex traits: From polygenic to omnigenic. Cell 169, 1177–1186 (2017).

39. Jagadeesh, K. A. et al. Identifying disease-critical cell types and cellular processes by integrating single-cell RNA-sequencing and human genetics. Nature Genetics 54, 1479–1492 (2022).

40. Maurano, M. T. et al. Systematic localization of common disease-associated variation in regulatory DNA. Science 337, 1190–1195 (2012).

41. Nathan, A., Baglaenko, Y., Fonseka, C. Y., Beynor, J. I. & Raychaudhuri, S. Multimodal single-cell approaches shed light on T cell heterogeneity. Current Opinion in Immunology 61, 17–25 (2019).

42. Trapnell, C. Defining cell types and states with single-cell genomics. Genome Research 25, 1491–1498 (2015).

43. Weinand, K. et al. (in preparation, 2023).

44. Korsunsky, I. et al. Fast, sensitive and accurate integration of single-cell data with Harmony. Nature Methods 16, 1289–1296 (2019).

45. Finucane, H. K. et al. Partitioning heritability by functional annotation using genome-wide association summary statistics. Nature Genetics 47, 1228–1235 (2015).

46. Kang, J. B. et al. Efficient and precise single-cell reference atlas mapping with Symphony. Nature Communications 12, 5890 (2021).

47. Mitsialis, V. et al. Single-cell analyses of colon and blood reveal distinct immune cell signatures of ulcerative colitis and crohn’s disease. Gastroenterology 159,591-608 (2020).

48. Gellatly, K. J. et al. ScRNA-seq of human vitiligo reveals complex networks of subclinical immune activation and a role for CCR5 in T-reg function. Science Translational Medicine 13, 610 (2021).

49. Zhang, F. et al. Cellular deconstruction of inflamed synovium defines diverse inflammatory phenotypes in rheumatoid arthritis. bioRxiv (2022).

50. Yang, S.-K. et al. Genome-wide association study of crohn’s disease in Koreans revealed three new susceptibility loci and common attributes of genetic susceptibility across ethnic populations. Gut 63, 80–87 (2013).

51. Jostins, L. et al. Host–microbe interactions have shaped the genetic architecture of inflammatory bowel disease. Nature 491, 119–124 (2012).

52. Liu, J. Z. et al. Association analyses identify 38 susceptibility loci for inflammatory bowel disease and highlight shared genetic risk across populations. Nature Genetics 47, 979–986 (2015).

53. Donlin, L. T. et al. Methods for high-dimensional analysis of cells dissociated from cryopreserved synovial tissue. Arthritis Research & Therapy 20, (2018).

54. Corces, M. R. et al. An improved ATAC-seq protocol reduces background and enables interrogation of frozen tissues. Nature Methods 14, 959–962 (2017).

55. Amemiya, H. M., Kundaje, A. & Boyle, A. P. The encode blacklist: Identification of problematic regions of the genome. Scientific Reports 9, (2019).

