## Supplementary Figures for "Dynamic regulatory elements in single-cell multimodal data capture autoimmune disease heritability and implicate key immune cell states"

**Gupta et al.**

**Description of Supplementary Tables**

**1-5.** Poisson GLM regression results across accessible peaks in B cells, T cells, myeloid cells, fibroblasts, and endothelial cells, respectively.

**6.** Enriched Gene Ontology processes for cell-state-invariant peaks.

**7-18.** Enriched Gene Ontology processes for each of the 12 dynamic peak clusters, from Cluster 0 through Cluster 11.

**19.** Description of the GWAS summary statistics data for the 19 traits analyzed for heritability estimates in this study.

**20.** Heritability enrichment and $\tau^*$ results for cell type annotations. Accessible ("OPEN") peaks in each of the five cell types.

**21.** Heritability enrichment and $\tau^*$ results for cell type annotations. Dynamic and cell-state-invariant peaks in each of the five cell types.

**22.** Heritability enrichment and $\tau^*$ results for T cell state annotations.

**23.** Meta-$\tau^*$ results for T cell state annotations across autoimmune diseases.

**24.** P-values for individual diseases for the discrete T and myeloid cell states. Values are included for both independent runs and those conditional on high-scoring cell states.

**25.** Heritability enrichment and $\tau^*$ results for myeloid cell state annotations.

**26.** Meta-$\tau^*$ results for myeloid cell state annotations across autoimmune diseases.

### Supplementary Figures

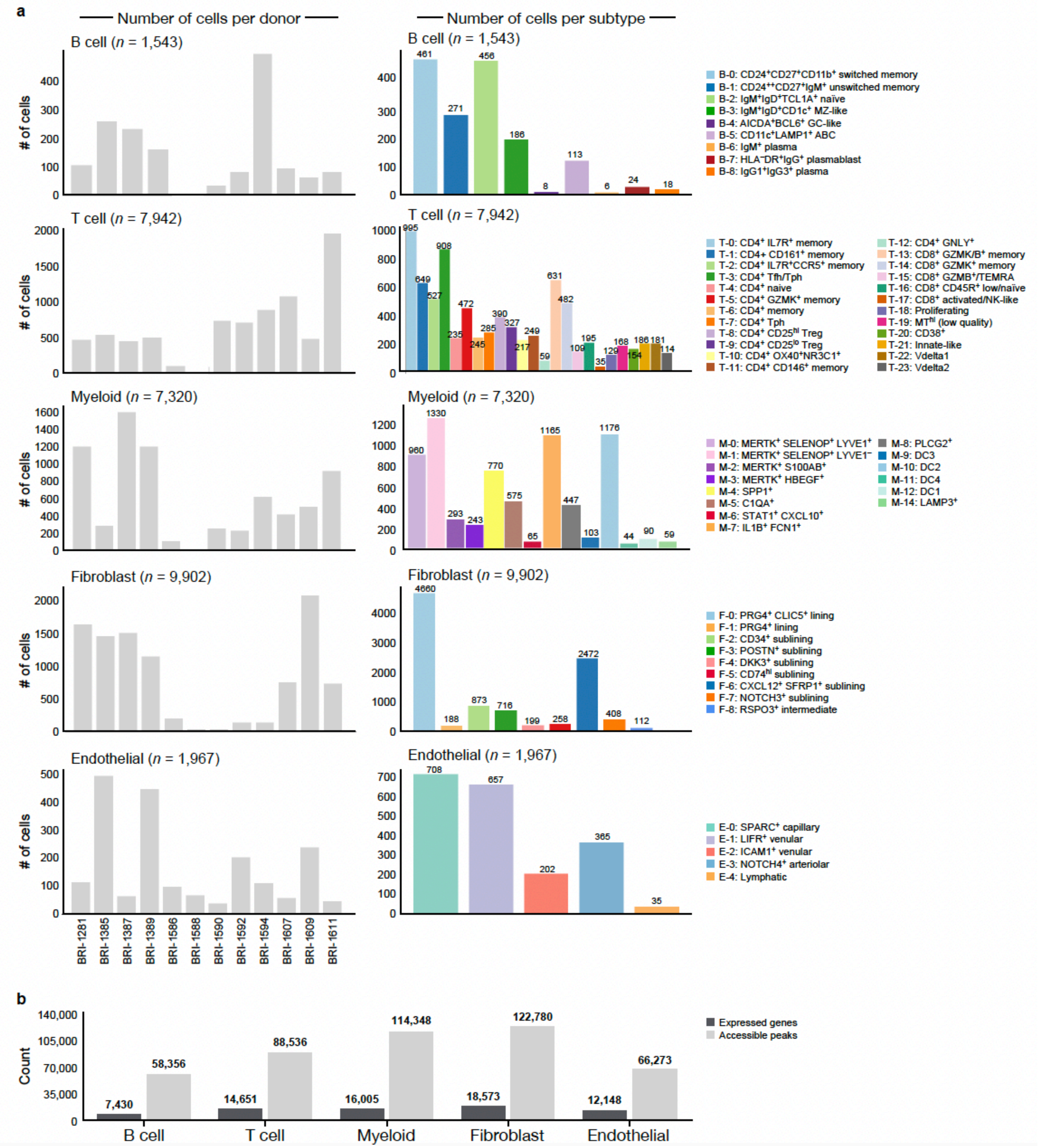


**Supplementary Fig. 1 | Number of cells per sample and per cell state. a,** Number of cells per donor and per cell subtype (discrete cell state) for each of the five cell types studied. Donor BRI-1281 has Osteoarthritis, while the other 11 donors have Rheumatoid Arthritis. **b,** Number of accessible peaks (open in at least 50 cells) and expressed genes (expressed in at least 50 cells) in each of the five cell types studied.

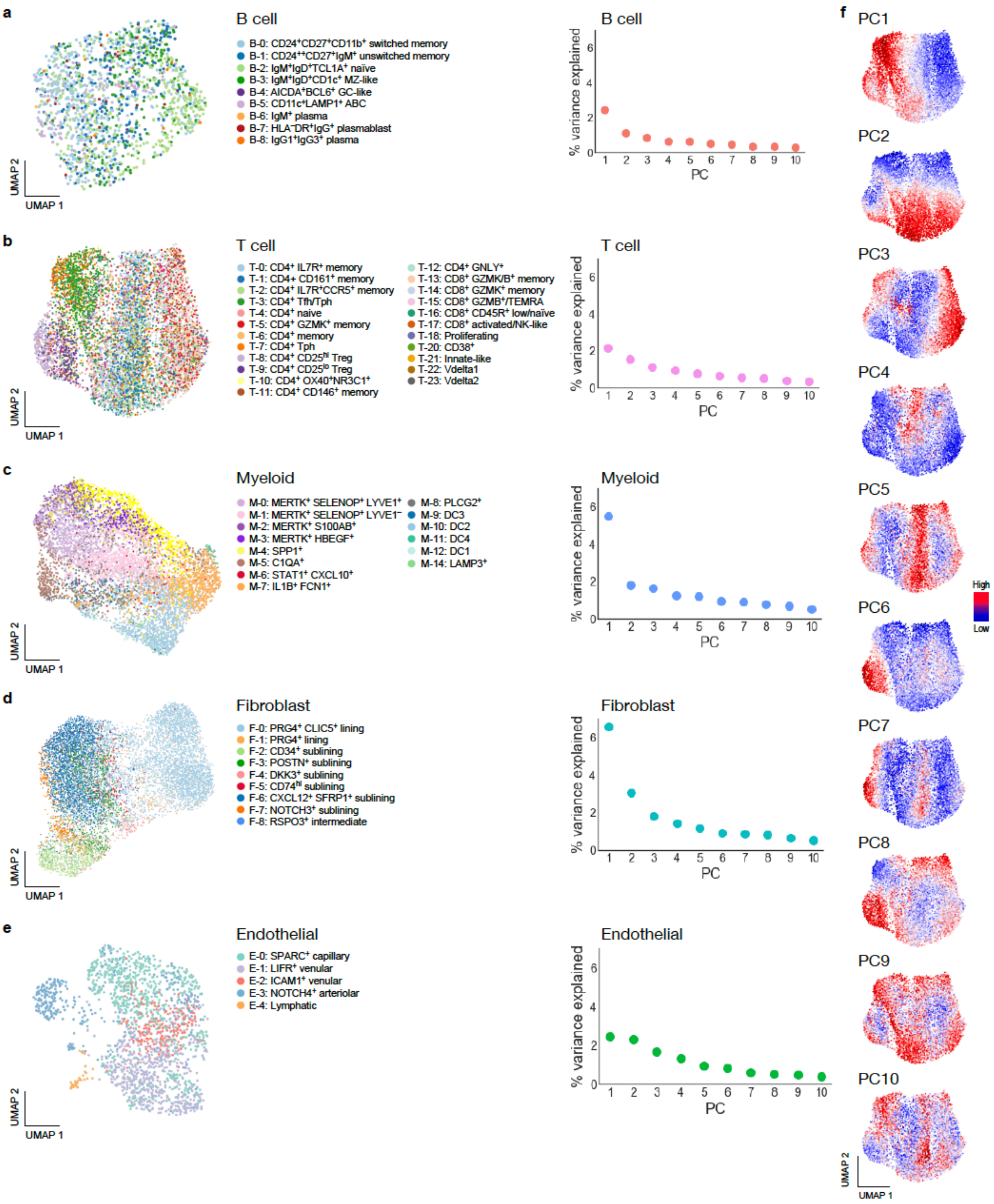


**Supplementary Fig. 2 | Discrete and continuous cell state attributes for each of the five cell types studied.** Post-processing. **a-e (left),** snRNA-seq-based UMAPs of B, T, myeloid, fibroblast, and endothelial discrete cell states, respectively. **a-e (right),** percent RNA variance explained by each of the top 10 RNA-PCs (B, T, myeloid, fibroblast, and endothelial cells, respectively). **f,** Top ten T cell RNA-PCs, with cells shaded by their PC loading. T cells shown as a case study of RNA-defined continuous cell states.

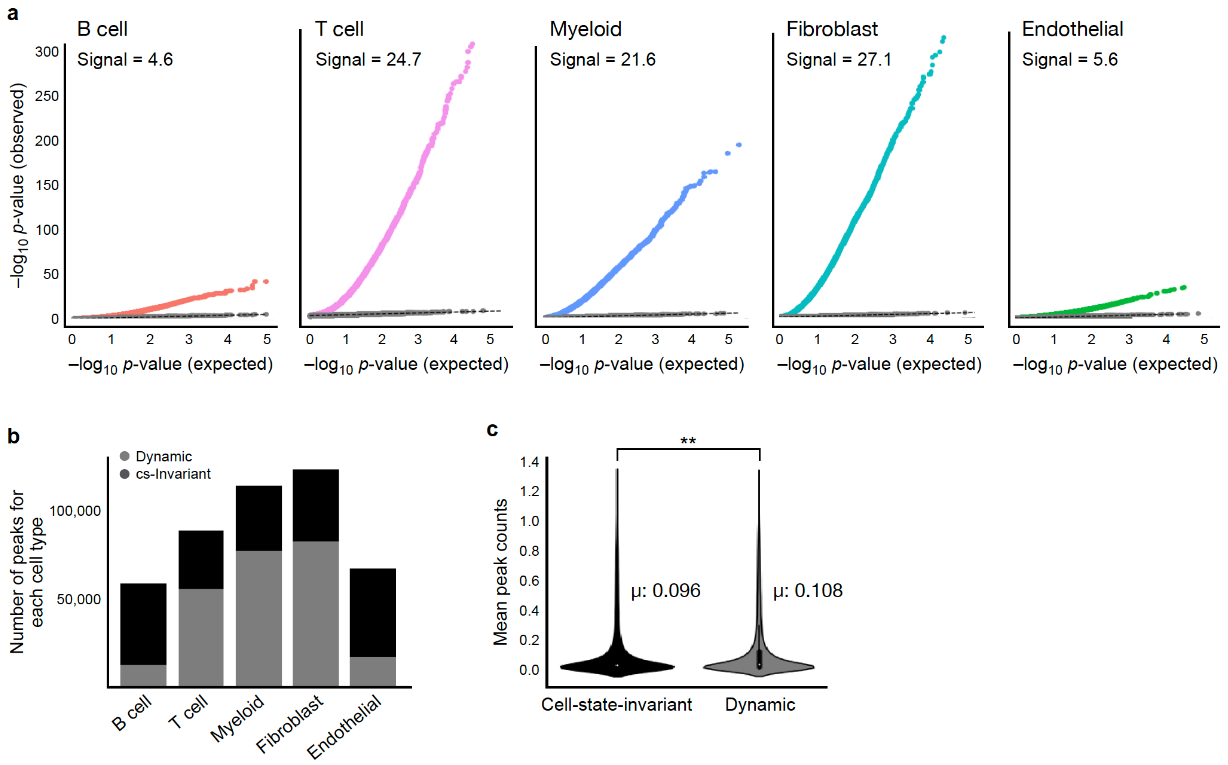


**Supplementary Fig. 3 | Poisson GLM regression QC checks and number of dynamic versus cell-state-invariant peaks, for the five cell types studied. a,** Quantile-quantile plot of expected versus observed p-values in the actual, observed data (colored by cell type) and null data (grey; generated by permuting the cell-PC assignments; inflation≤1.1 for all five cell types' null p-values), relative to the expected p-value distributions (uniform distribution). This Q-Q plot is shown for each of the five cell types studied. The y=x line is shown as a dashed black line. **b,** Number of accessible but cell-state-invariant accessible (black) versus dynamic (grey) peaks in each cell type. The ratio of grey to the total value of black bars indicates the fraction of accessible peaks in each cell type that are also dynamic in that cell type. **c,** Distribution of mean peak counts for cell-state-invariant and dynamic peaks (in T cells, as a representative example). We used a two-sided Wilcoxon rank sum test to determine that the two distributions are distinct (in T cells, $\mu_{dynamic}$=0.108 versus $\mu_{cs\text{-}invariant}$=0.096, p<1e-20), suggesting that power differences are present but play a minimal role.

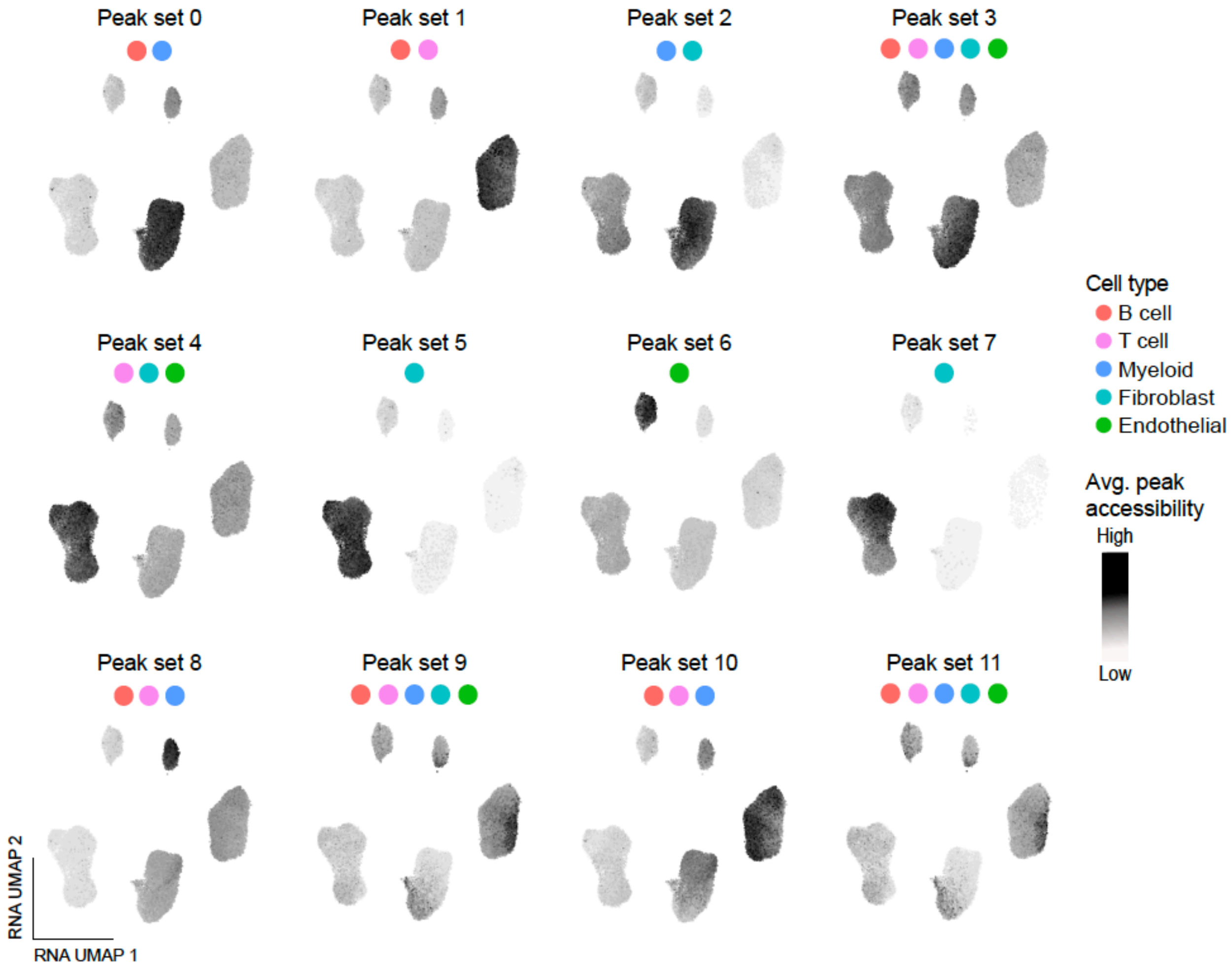


**Supplementary Fig. 4 | Cell types represented by each dynamic peak set.** In this snRNA-based UMAP of all five cell types analyzed, each dot represents a cell. Shading reflects the mean accessibility of the peaks in each cluster (0-11). For each peak set, the cell types with relatively higher mean peak accessibility are circled with their respective color. Supplementary Tables 7-18 have the top Gene Ontology biological processes enriched by these peak sets (peaks were assigned to the gene whose promoter region (TSS +/- 1kb) they overlapped with, and the gene set corresponding to each peak set was run through Gene Ontology gene set enrichment).

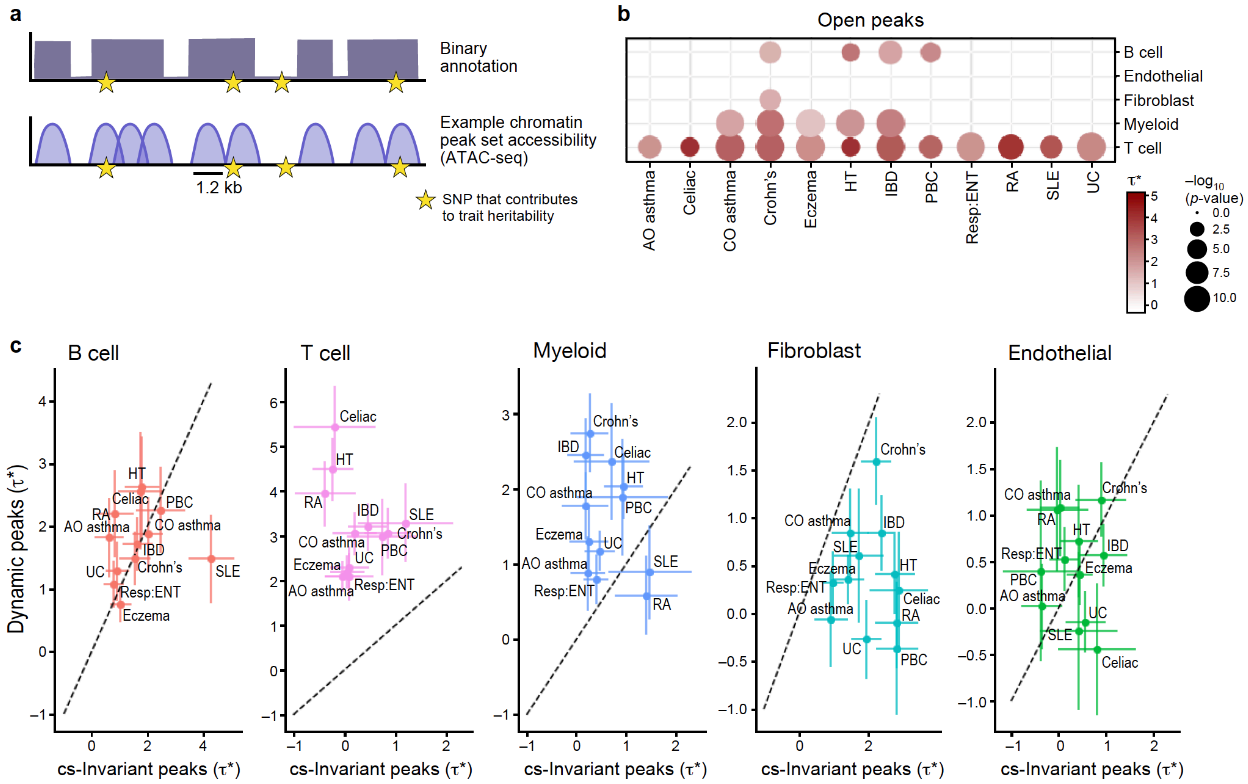


**Supplementary Fig. 5 | Per-SNP effect sizes across annotations for the five cell types. a,** Genome annotation strategy for a toy peak set, to create input into S-LDSC. Each peak was 1.2 kb in size. **b,** Open peaks' $\tau^*$ within each cell type, across the 12 autoimmune conditions studied. **c,** $\tau^*$ for dynamic versus cell-state-invariant ("cs-invariant") peaks within each cell type (error bars indicate 95% CI). Dotted line indicates y=x. Trait acronyms: adult-onset asthma: Asthma (a), child-onset asthma: Asthma (c), Crohn's disease: Crohn's, hypothyroidism: HT, inflammatory bowel disease: IBD, primary biliary cirrhosis: PBC, respiratory ear-nose-throat disease: Resp:ENT, rheumatoid arthritis: RA, systemic lupus erythematosus: SLE, ulcerative colitis: UC.

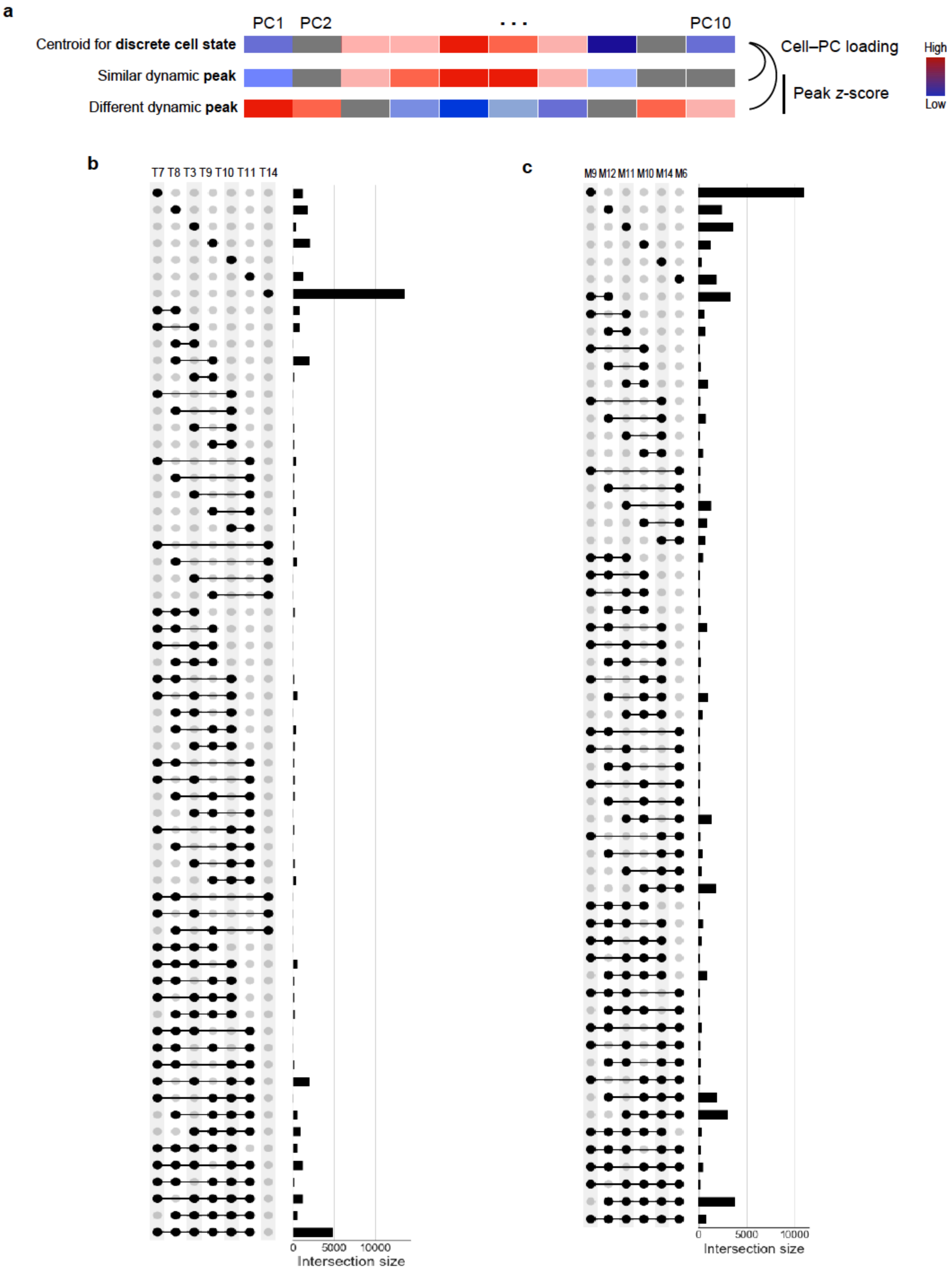
a
PC1
PC2
...
PC10
Centroid for discrete cell state
Similar dynamic peak
Different dynamic peak
Cell–PC loading
Peak z-score
High
Low
b
T7 T8 T3 T9 T10 T11 T14
0
5000
10000
Intersection size
c
M9 M12 M11 M10 M14 M6
0
5000
10000
Intersection size

**Supplementary Fig. 6 | Defining and characterizing the heritability captured by T and myeloid discrete cell states. a,** Strategy for determining the most similar 25% of peaks for each discrete cell state. For the relevant cell type, we determined the centroid for each RNA PC across all cells of a given state and calculated the cosine similarity between these values and each dynamic peak's z-scores for these 10 PCs from the Poisson GLM regression. **b,** Heatmap of the number of peaks either unique or shared in the top 25% most similar set across all pairs of discrete T cell states. **c,** Heatmap of the number of peaks either unique or shared in the top 25% most similar set across all pairs of discrete myeloid cell states.

| | T-0 | T-1 | T-2 | T-3 | T-4 | T-5 | T-6 | T-7 | T-8 | T-9 | T-10 | T-11 | T-12 | T-13 | T-14 | T-15 | T-16 | T-17 | T-18 | T-20 | T-21 | T-22 | T-23 |
|---|---|---|---|---|---|---|---|---|---|---|---|---|---|---|---|---|---|---|---|---|---|---|---|
| T-0 | | | | | | | | | | | | | | | | | | | | | | | |
| T-1 | 11796 | | | | | | | | | | | | | | | | | | | | | | |
| T-2 | 10064 | 11121 | | | | | | | | | | | | | | | | | | | | | |
| T-3 | 1864 | 1546 | 235 | | | | | | | | | | | | | | | | | | | | |
| T-4 | 8695 | 7149 | 5185 | 4341 | | | | | | | | | | | | | | | | | | | |
| T-5 | 825 | 1171 | 3083 | 54 | 321 | | | | | | | | | | | | | | | | | | |
| T-6 | 5489 | 4666 | 4590 | 540 | 5757 | 4148 | | | | | | | | | | | | | | | | | |
| T-7 | 706 | 896 | 159 | 11483 | 2367 | 381 | 223 | | | | | | | | | | | | | | | | |
| T-8 | 1399 | 1336 | 581 | 7805 | 4344 | 822 | 1948 | 8246 | | | | | | | | | | | | | | | |
| T-9 | 4053 | 3217 | 1525 | 8342 | 8362 | 130 | 2649 | 6609 | 9129 | | | | | | | | | | | | | | |
| T-10 | 2494 | 2096 | 501 | 12482 | 5219 | 0 | 630 | 10736 | 8733 | 9610 | | | | | | | | | | | | | |
| T-11 | 3654 | 3743 | 1745 | 10792 | 5306 | 0 | 501 | 9710 | 7497 | 8600 | 11481 | | | | | | | | | | | | |
| T-12 | 5803 | 6245 | 8379 | 0 | 2801 | 7607 | 6288 | 0 | 197 | 305 | 0 | 17 | | | | | | | | | | | |
| T-13 | 1206 | 1733 | 3971 | 14 | 107 | 12189 | 2700 | 407 | 642 | 26 | 0 | 0 | 7925 | | | | | | | | | | |
| T-14 | 1822 | 2279 | 4682 | 4 | 346 | 11548 | 2702 | 138 | 460 | 59 | 0 | 0 | 8482 | 12989 | | | | | | | | | |
| T-15 | 2759 | 3465 | 5986 | 0 | 644 | 9797 | 2194 | 28 | 452 | 92 | 0 | 0 | 8921 | 11319 | 12248 | | | | | | | | |
| T-16 | 7900 | 8641 | 11155 | 0 | 3946 | 4911 | 3677 | 0 | 248 | 831 | 2 | 423 | 9542 | 5926 | 6844 | 8294 | | | | | | | |
| T-17 | 2938 | 3739 | 6302 | 0 | 513 | 9453 | 2010 | 65 | 291 | 11 | 0 | 0 | 8706 | 10974 | 11705 | 12509 | 8529 | | | | | | |
| T-18 | 66 | 604 | 2115 | 1049 | 0 | 9911 | 1113 | 2610 | 2187 | 118 | 572 | 294 | 4999 | 10545 | 9631 | 8664 | 3647 | 8710 | | | | | |
| T-20 | 1398 | 1994 | 4254 | 35 | 205 | 10521 | 1750 | 365 | 701 | 83 | 0 | 0 | 7265 | 11815 | 12036 | 11389 | 6554 | 11450 | 9898 | | | | |
| T-21 | 7674 | 9709 | 9644 | 940 | 4343 | 3480 | 4131 | 946 | 1168 | 1730 | 1087 | 2567 | 7660 | 4020 | 4463 | 5462 | 8848 | 5772 | 2919 | 4141 | | | |
| T-22 | 1009 | 1562 | 3697 | 97 | 197 | 11974 | 2353 | 536 | 954 | 151 | 4 | 0 | 7516 | 12957 | 12758 | 11366 | 5822 | 10806 | 10548 | 11909 | 4038 | | |
| T-23 | 12251 | 11074 | 9052 | 2371 | 9664 | 723 | 5218 | 871 | 1691 | 4849 | 2968 | 4023 | 5292 | 913 | 1525 | 2443 | 7404 | 2567 | 15 | 1189 | 7697 | 929 | |

**Supplementary Fig. 7 | Number of dynamic peaks that overlap in the top 25% most similar peak set for each discrete T cell state.** Similar peaks determined by cosine similarity between each peak's regression Z-scores across the top 10 RNA PCs and each cell state centroid's PC positions for the top 10 RNA PCs.

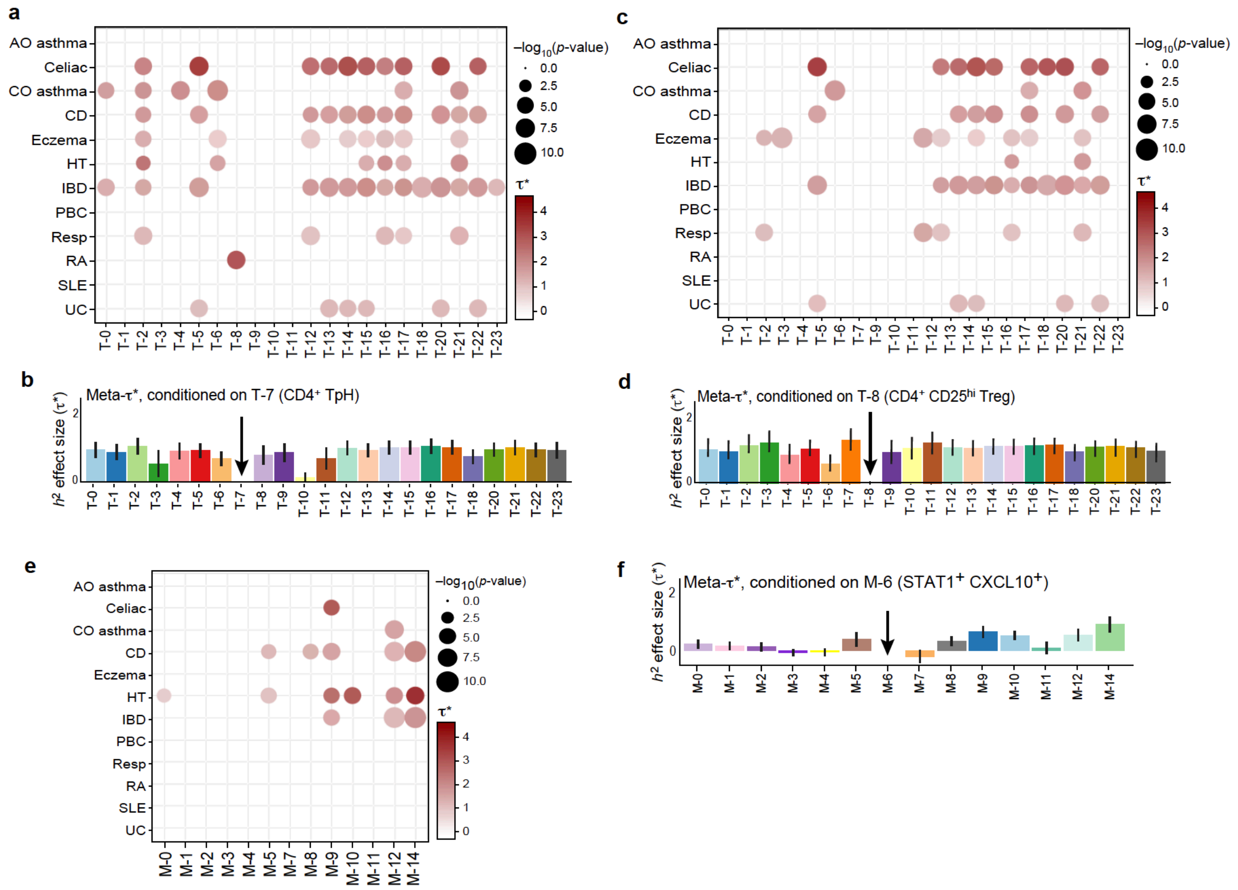


**Supplementary Fig. 8 | Discrete T and myeloid cell state-based individual and meta conditional $\tau^*$ values in autoimmunity, using the top 25% of similar peaks to represent each cell state. a,** Individual tau $\tau^*$ values for all T cell states, conditioned on T-7. All values shown have p<0.001. **b,** Meta-tau $\tau^*$ value across all T cell states, conditioned on T-7. Error bars indicate +/- 95% CIs. **c,** Individual tau $\tau^*$ values for all T cell states, conditioned on T-8. All values shown have p<0.001. **d,** Meta-tau $\tau^*$ value across all T cell states, conditioned on T-8. Error bars indicate +/- 95% CIs. **e,** Individual tau $\tau^*$ values for all myeloid cell states, conditioned on M-6. All values shown have p<0.001. **f,** Meta-tau $\tau^*$ value across all myeloid cell states, conditioned on M-6. Error bars indicate +/- 95% CIs.

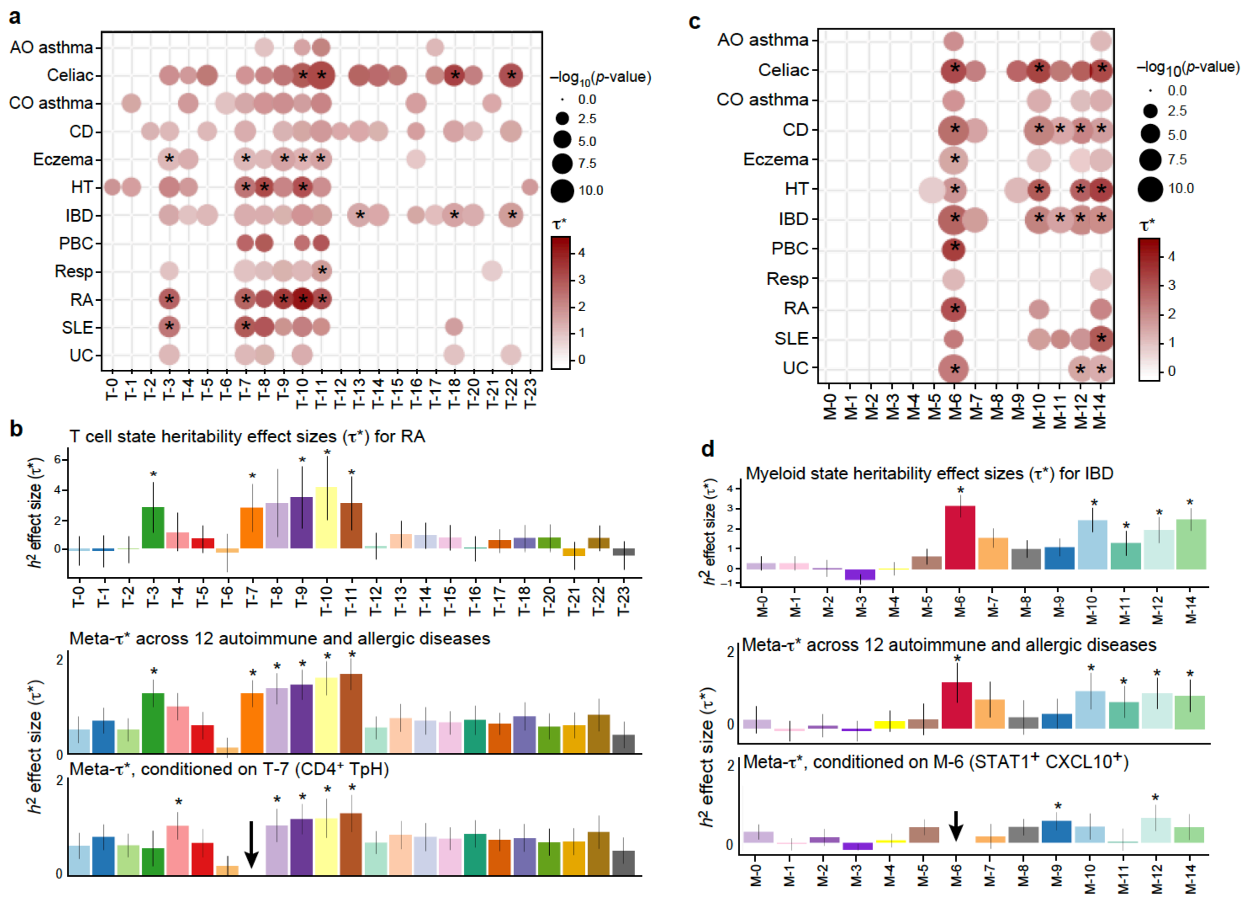


**Supplementary Fig. 9 | Discrete T and myeloid cell state-based individual and meta independent and conditional $\tau$* values in autoimmunity, using the top 10% of similar peaks to represent each cell state. a,** Individual $\tau$* values for each discrete T cell state in the 12 autoimmune and allergic conditions analyzed, using the top 10% of peaks for each state's annotation. Only nominally significant (p<0.05) annotations are shown, and those that remain significant upon Bonferroni correction (p<0.05/12) are marked with an asterisk. **b,** (top) Annotation effect size ($\tau$*) for each discrete T cell state, for rheumatoid arthritis (RA). Error bars indicate +/- 95% CIs, and asterisks indicate p < 0.05/12. Meta (middle) independent and (bottom) T-7 conditional $\tau$* values for T cell states, using the top 10% of peaks for each state's annotation. Asterisks for RA indicate p<0.05/12 and for meta-analyses indicate p<1e-30.
**c,** Individual $\tau$* values for each discrete myeloid cell state in the 12 autoimmune and allergic conditions analyzed, using the top 10% of peaks for each state's annotation. Only nominally significant (p<0.05) annotations are shown, and those that remain significant upon Bonferroni correction (p<0.05/12) are marked with an asterisk. **d,** (top) Annotation effect size ($\tau$*) for each discrete myeloid cell state, for inflammatory bowel disease (IBD). Error bars indicate +/- 95% CIs, and asterisks indicate p < 0.05/12. Meta (middle) independent and (bottom) M-6 conditional $\tau$* values for myeloid cell states, using the top 10% of peaks for each state's annotation. Asterisks for IBD indicate p<0.05/12 and for meta-analyses indicate p<1e-15.

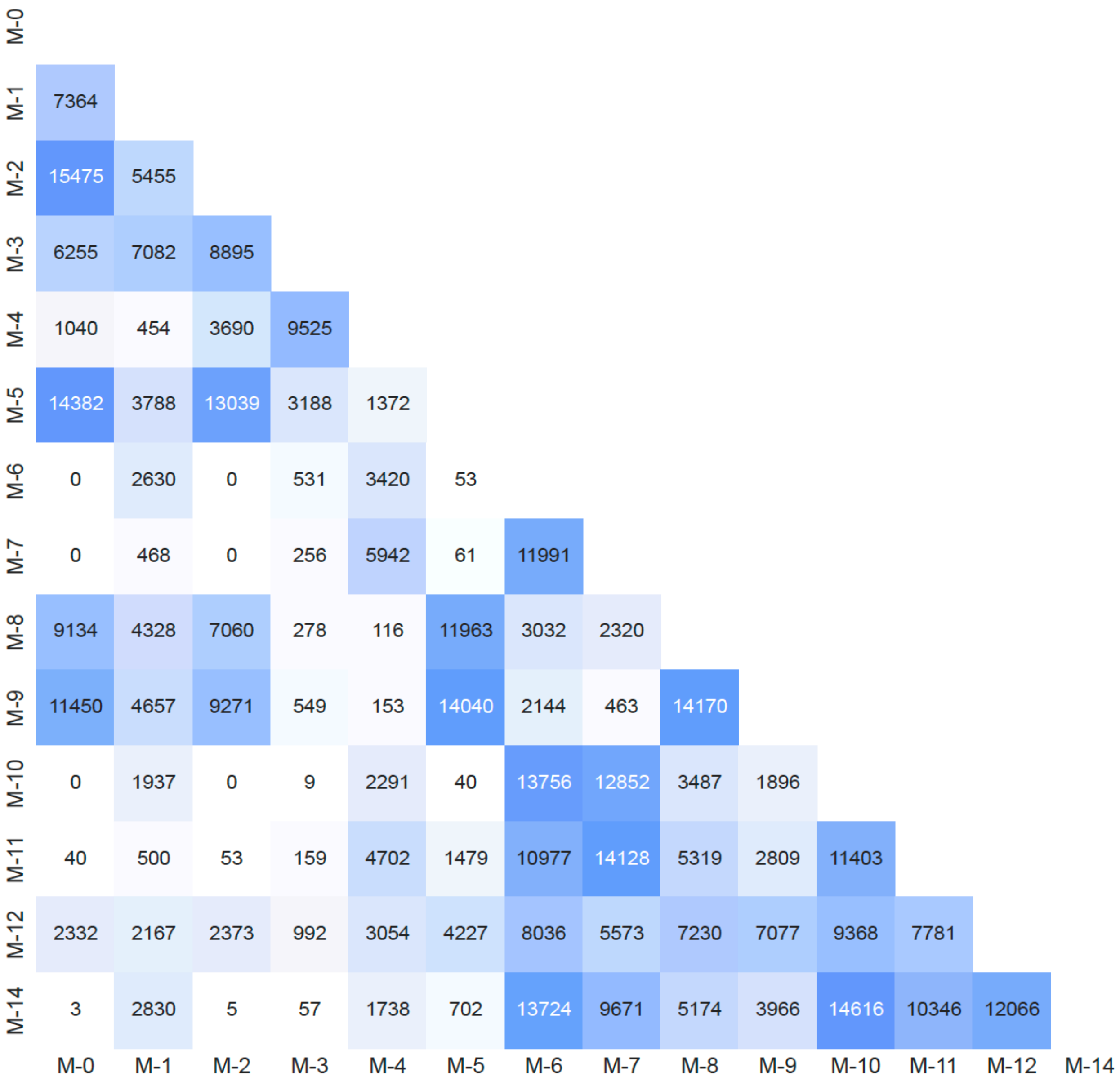


**Supplementary Fig. 10 | Number of dynamic peaks that overlap in the top 25% most similar peak set for each discrete myeloid cell state.** Similar peaks determined by cosine similarity between each peak's regression Z-scores across the top 10 RNA PCs and each cell state centroid's PC positions for the top 10 RNA PCs.
